# Systematic evaluation of medication adherence determinants across 137 ingredients on population-level real-world health data

**DOI:** 10.1101/2025.02.10.25319884

**Authors:** Kerli Mooses, Marek Oja, Maria Malk, Helene Loorents, Maarja Pajusalu, Nikita Umov, Sirli Tamm, Johannes Holm, Hanna Keidong, Taavi Tillmann, Sulev Reisberg, Jaak Vilo, Raivo Kolde

## Abstract

The current knowledge about medication adherence (MA) is based on studies focusing only on few health conditions. We utilised a representative dataset with electronic health records, claims, and prescribed medications to 1) evaluate the effect of multiple factors affecting MA in a consistent manner across 137 ingredients, and 2) calculate individual medication adherence score (IMAS), evaluate its predictive power, stability over time, and impact on health outcomes. The MA ranged from 0.423 (albuterol, 95% CI 0.414–0.432) to 0.922 (warfarin, 95% CI 0.917–0.926). The demographic, health- and medication-related factors explained 11.6% and IMAS 22% of the variation in adherence. IMAS predicted adherence across medication classes, reduced the risk of overall hospitalisation (Hazard ratio = 0.76, 95% CI 0.60–0.97, p<0.05) and cause-specific incidence for 17 conditions. Thus, medication-taking behaviour represents a broader patient-level phenomenon manifesting consistently across medications, suggesting its potential for personalised interventions in clinical practice and more efficient public health strategies and policies.

## Introduction

Adherence to medications is crucial for achieving better health and clinical outcomes,^1–3^ which in turn reduces hospitalisation and annual healthcare costs.^3,4^ Moreover, a 16–21% reduction in mortality risk has been reported for patients with good adherence.^1,5^ Medication adherence is a complex multifactorial phenomenon.^6,7^ A review of systematic reviews identified more than 800 individual, medication-related and healthcare system-related factors potentially influencing medication adherence.^7^ However, the effect size, significance and even direction were found to be inconsistent across studies.^7^ Thus, more research is needed on factors influencing medication adherence.

One challenge of synthesising the existing knowledge lies in the nature of studies that mostly focus on one or a few selected chronic conditions.^1,4,8^ As a result, little is known about the adherence patterns within the same individual for co-prescribed medications. Considering that the prevalence of polypharmacy is 55% among people older than 65 years,^9^ broadening the analysis with a wide range of medications for chronic conditions is necessary. Moreover, to date, there is a gap in understanding the extent to which overall adherence is shaped by medication-specific factors, as opposed to person-specific traits or different life events. Therefore, a qualitatively different approach is needed to disentangle the complex network of clinical, socio-economic and behavioural factors influencing medication adherence.

The emergence of population-level prescription databases, linked with patient medical records, allows to systematically and with consistent methodology evaluate the influence of different factors on medication adherence while distinguishing medication-level, person-specific and time-dependent effects. In the current study, we take advantage of a representative random sample of the Estonian population to study the determinants of medication adherence over a hundred ingredients prescribed for long-term use. As a novel approach, we calculate individual medication adherence score (IMAS) using a wide range of ingredients prescribed for chronic conditions and apply IMAS in further analysis. More precisely, we first compare the effect sizes of multiple factors affecting medication adherence calculated in a consistent manner across 137 ingredients. Secondly, we calculate IMAS using information about 137 ingredients, describe its predictive power and stability over time, and evaluate its effect on health outcomes.

## Results

### Medication adherence

Less than half of the subjects in the database (43.0%, N = 64 837) had at least one ingredient under observation prescribed twice within a year and it was possible to calculate CMA. That group of patients included 57% females with an average age of 56.5 ± 21.8 years (Table 1).

**Table 1.** Descriptives of 64 837 subjects included in the analysis.

| Characteristics | N = 64 837 |
| --- | --- |
| Gender, N (%) |  |
| Female | 37,111 (57.2%) |
| Male | 27,726 (42.8%) |
| Body mass index category, N (%) |  |
| Obese | 10,690 (16.5%) |
| Overweight | 9,396 (14.5%) |
| Under or normal weight | 8,003 (12.3%) |
| Unknown | 36,748 (56.7%) |
| Mean age in years (sd) | 56.50 (21.75) |
| Mean CMA (sd) | 0.75 (0.21) |

On average 75% of the days were covered with prescribed medication (Table 1). There were 52.1% of subjects with an average CMA over 0.8 and 7.7% with an average CMA below 0.4. Overall, females had significantly higher average CMA compared with males (0.76, 95% CI 0.76–0.77 vs 0.73, 95% CI 0.72– 0.74 respectively, p < 0.001). At least one comorbidity from the Charlson Comorbidity Index was present for 64.9%, and depression was diagnosed for 21.7% of subjects.

When looking at the raw CMA values, the subjects took on average 4.3 ± 3.7 different ingredients during the study period. The CMA per ingredient ranged from 0.423 (albuterol, 95% CI 0.414–0.432) to 0.922 (warfarin, 95% CI 0.917–0.926) (Supplementary Table 1). The ingredients were assigned to a disease groups based on the most prevalent diagnoses on their prescription (Supplementary Table 2). Eight disease groups had an average CMA of 0.8 or higher – disorders of thyroid gland (E00–E07), presence of cardiac and vascular implants and grafts (Z95), malignant neoplasm of breast (C50), Parkinson disease (G20), arrhythmias (I46–I49), malignant neoplasm of prostate (C61), osteoporosis (M80, M81), and Glaucoma (H40-H42) (Table 2). The lowest medication adherence was for diseases of the digestive system (K00–K93) (CMA = 0.488, 95% CI 0.479–0.497). The highest proportion of adherent subjects (CMA ≥ 0.8) was for malignant neoplasm of breast (82.0%) and disorders of thyroid gland (78.4%), while the lowest proportion of adherent subjects was among diseases of respiratory system (22.7%) and digestive system (24.3%) (Table 2).

**Table 2.**
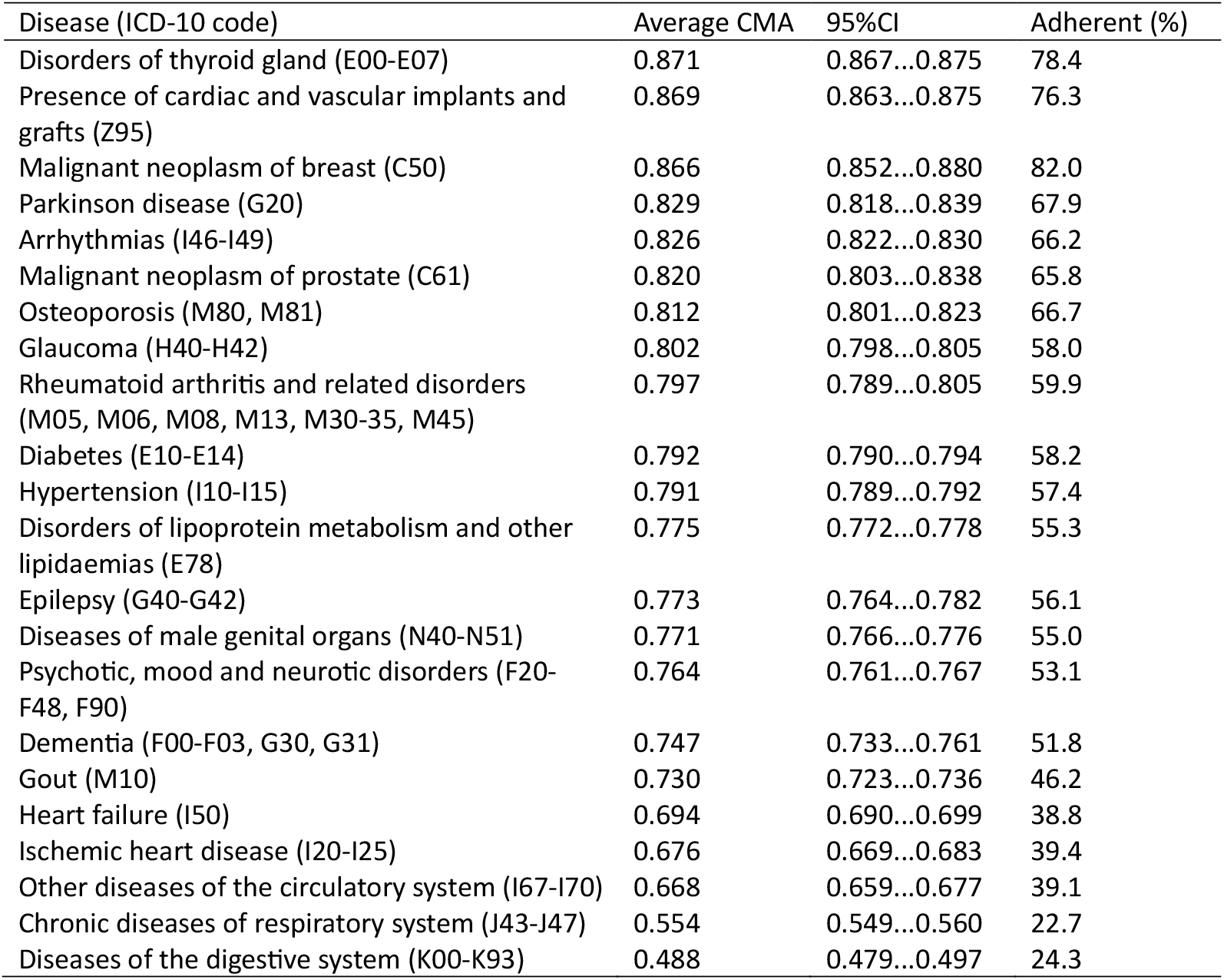
Average CMA with 95% confidence intervals and proportion of adherent persons (CMA ≥ 0.8) by disease groups.

| Disease (ICD-10 code) | Average CMA | 95%CI | Adherent (%) |
| --- | --- | --- | --- |
| Disorders of thyroid gland (E00-E07) | 0.871 | 0.867...0.875 | 78.4 |
| Presence of cardiac and vascular implants and grafts (Z95) | 0.869 | 0.863...0.875 | 76.3 |
| Malignant neoplasm of breast (C50) | 0.866 | 0.852...0.880 | 82.0 |
| Parkinson disease (G20) | 0.829 | 0.818...0.839 | 67.9 |
| Arrhythmias (I46-I49) | 0.826 | 0.822...0.830 | 66.2 |
| Malignant neoplasm of prostate (C61) | 0.820 | 0.803...0.838 | 65.8 |
| Osteoporosis (M80, M81) | 0.812 | 0.801...0.823 | 66.7 |
| Glaucoma (H40-H42) | 0.802 | 0.798...0.805 | 58.0 |
| Rheumatoid arthritis and related disorders (M05, M06, M08, M13, M30-35, M45) | 0.797 | 0.789...0.805 | 59.9 |
| Diabetes (E10-E14) | 0.792 | 0.790...0.794 | 58.2 |
| Hypertension (I10-I15) | 0.791 | 0.789...0.792 | 57.4 |
| Disorders of lipoprotein metabolism and other lipidaemias (E78) | 0.775 | 0.772...0.778 | 55.3 |
| Epilepsy (G40-G42) | 0.773 | 0.764...0.782 | 56.1 |
| Diseases of male genital organs (N40-N51) | 0.771 | 0.766...0.776 | 55.0 |
| Psychotic, mood and neurotic disorders (F20-F48, F90) | 0.764 | 0.761...0.767 | 53.1 |
| Dementia (F00-F03, G30, G31) | 0.747 | 0.733...0.761 | 51.8 |
| Gout (M10) | 0.730 | 0.723...0.736 | 46.2 |
| Heart failure (I50) | 0.694 | 0.690...0.699 | 38.8 |
| Ischemic heart disease (I20-I25) | 0.676 | 0.669...0.683 | 39.4 |
| Other diseases of the circulatory system (I67-I70) | 0.668 | 0.659...0.677 | 39.1 |
| Chronic diseases of respiratory system (J43-J47) | 0.554 | 0.549...0.560 | 22.7 |
| Diseases of the digestive system (K00-K93) | 0.488 | 0.479...0.497 | 24.3 |

Methylphenidate users were the youngest (13.6 years, 95% CI 12.9–14.3), while donepezil and isosorbide users were the oldest (77.4 years, 95% CI 76.5–78.3 and 77.3 years, 95% CI 76.9–77.7 respectively) (Supplementary Table 3).

### Factors influencing medication adherence

The CMA variation was modelled using linear mixed models (LMM), including several demographic, health- and medication-related variables as fixed effects, as well as random person-level effects, capturing the correlation between the observed adherence values across medications and time periods for the same person. The whole model described 33.6% of the variation in medication adherence measurements, while 22.0% of that was explained by the person-specific effects. From demographic variables, gender had no effect on medication adherence, while 80 years and older people had higher (0.11, 95% CI 0.10–0.13) medication adherence compared to those younger than 20 years (Fig. 1, Supplementary Table 4). Several health-related factors like the occurrence of depression, dementia, hospitalisation or comorbidities did not have an effect on medication adherence. The expected CMA values were more than 0.1 units lower for 27 and higher for two medications compared to metoprolol which was used as the comparator in the analysis. Some diagnoses were associated with adherence. However, very few like malignant neoplasm of breast (C50) (0.30, 95% CI 0.26–0.34) and glaucoma (H40) (0.35, 95% CI 0.21–0.49) displayed comparably large effect sizes to medication adherence.

**Fig. 1.**
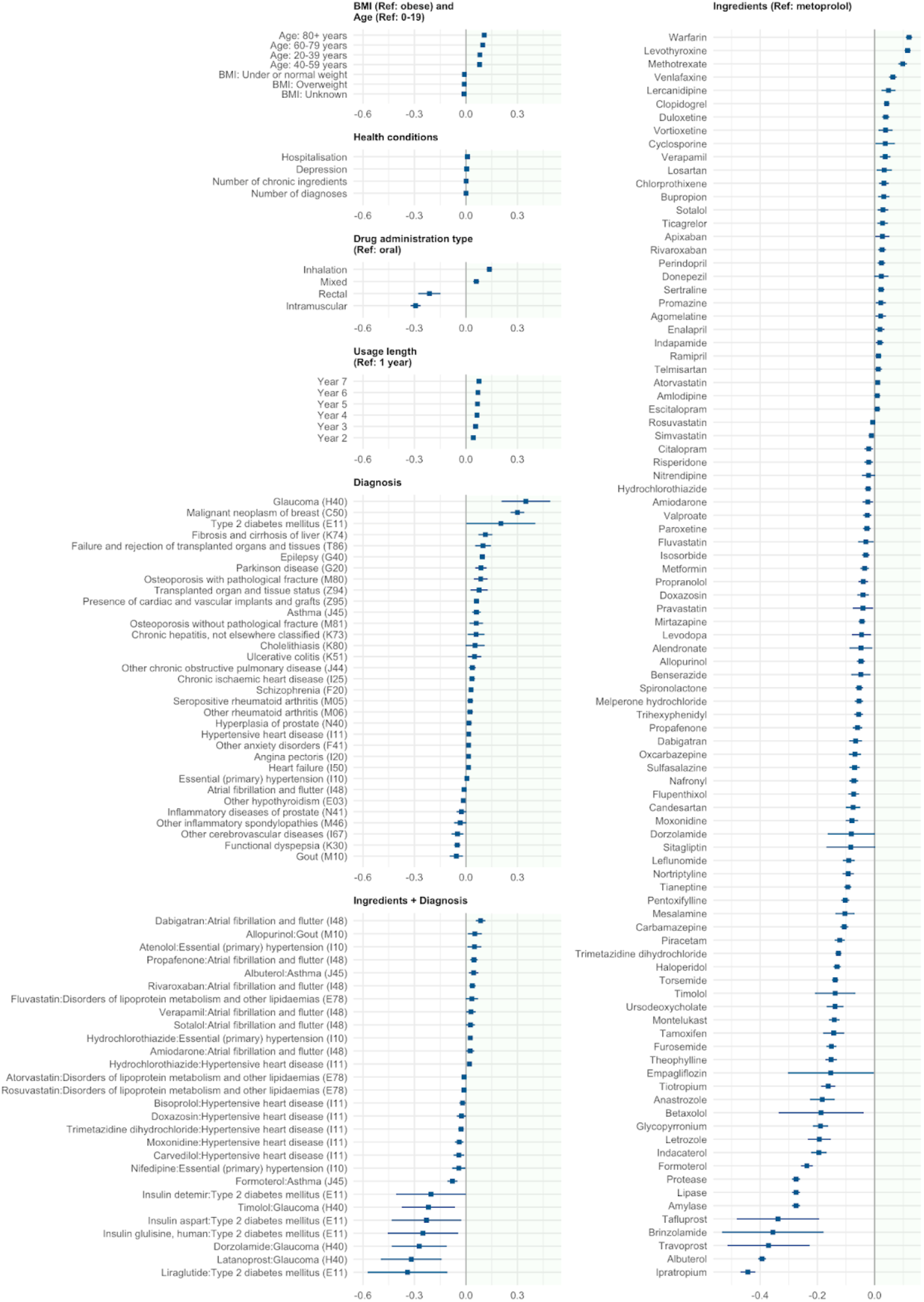
Variables influencing medication adherence based on linear mixed model analysis. The effect estimates and 95% confidence intervals of the variables statistically significant after the Bonferroni correction are presented. Information for all included variables is presented in Supplementary Table 4.

### Analysis with IMAS

IMAS is the person-specific effect extracted from LMM including several demographic, health-and medication-related variables. It represents an individual’s inherent medication-taking behaviour – their personal “baseline” adherence tendency that persists across prescriptions and time, after accounting for demographic, medication- and health-related factors. IMAS was included in further analysis to evaluate its impact and continuity over time. First, the IMAS calculated for only ingredients prescribed for cardio-vascular diseases (CVD) was added into a LMM calculated for all other ingredients prescribed for chronic conditions (non-CVD). The inclusion of IMAS based on CVD ingredients significantly increased the medication adherence for non-CVD ingredients (estimate = 0.45, p < 0.001). Moreover, the R-squared of fixed effects was slightly higher for a model where IMAS from the CVD model was added as fixed effects compared to the model without this IMAS (R^2^ = 0.14 vs R^2^ = 0.11, respectively).

Next, the continuity of the IMAS over time was assessed. The correlation between yearly IMAS declined over time, with the highest correlation being between consecutive years (r_avg_ = 0.46, range 0.36 to 0.49, p < 0.05) (Fig. 2).

**Fig. 2.**
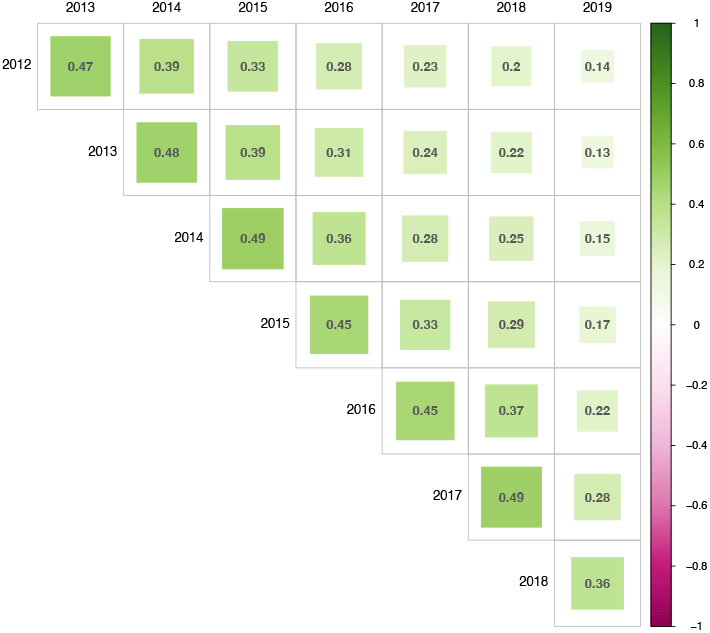
Correlation of IMAS from 2012 to 2019. All correlations are statistically significant (p < 0.05).

Last we sought to evaluate if IMAS predicts health outcomes in the future. For this purpose, IMAS calculated for the period 2012 to 2016 was used to predict the health outcomes in 2017–2019 using a Cox proportional hazards model. From 2017 to 2019, there were 5904 hospitalisations. We found that higher IMAS reduced the risk of overall hospitalisation (HR = 0.76, 95% CI 0.60–0.97, p < 0.05), but no effect on cause-specific hospitalisation risk after Bonferroni adjustment was detected (Supplementary Table 5).

We used the same approach to predict the incidence of a collection of chronic conditions. Out of 32 chronic conditions higher IMAS was associated with a lower risk of cause-specific incidence for 17 conditions after Bonferroni correction. The reduction in incidence risk ranged from 27% for atherosclerosis/PAOD and diseases of liver to 58% for diseases of stomach (Fig. 3, Supplementary Table 6).

**Fig. 3.**
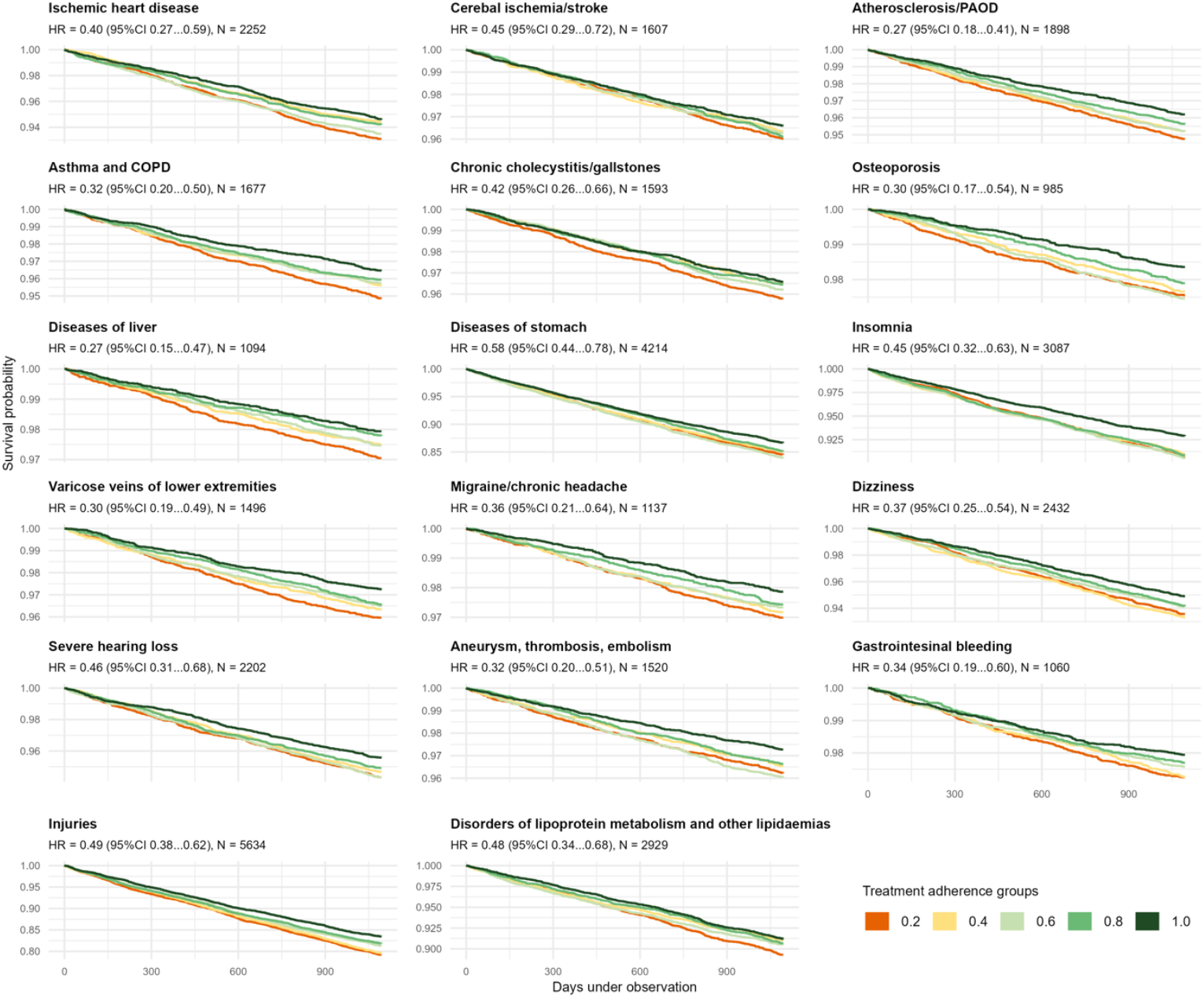
Kaplan-Meier curves for cause-specific incidence for statistically significant diagnoses.

## Discussion

This study set out to investigate medication adherence and its impact on health outcomes with a novel method incorporating more than 130 ingredients and a variety of demographic, health- and medication-related variables from real-world electronic health data. We modelled adherence values using LMM revealing low predictive power for many commonly studied risk factors, but consistent person-level behaviour across medications and time. The calculated IMAS was predictive for both future adherence as well as health outcomes.

Our extensive analysis across 137 ingredients revealed a significant variation in medication adherence based on ingredient and disease – there was more than a two-fold difference between ingredients with the best and the worst adherence and almost a four-fold difference in the prevalence of good adherence between diseases. The prevalence of patients with good adherence in our study was slightly lower for antidiabetic agents (58%) and similar for statins (55%) and antihypertensives (57%) compared with the previous meta-analysis.^10^ However, the diversity in adherence calculation methods and ingredient selection complicates the comparison between studies and diseases. Also, we expect differences in adherence patterns between localities and healthcare systems to be present, raising the need to conduct similar studies with consistent methodology on the growing number of population-based prescription databases worldwide.

It is widely accepted that medication adherence is influenced by several individual, medication-related and healthcare system related factors.^3,7,11^ Our results indicated that a considerable amount of observed variables had a statistically significant impact on medication adherence, but the effect size was very small. This finding is in accordance with previous studies where the observed effects were very small and often inconsistent between studies for most patient-related factors.^7,8^ More importantly, we found that almost a quarter of the medication adherence variance was explained by IMAS, capturing more variation than the fixed effects in the mixed effect model. The significance of IMAS has several implications for further medication adherence research.

First, the medication-taking behaviour represents a broader patient-level phenomenon that manifests consistently across therapeutic classes, and it should be studied as such. There is a need for a better understanding of medication adherence behaviour through larger studies on more ingredients in diverse patient cohorts across the world. Also, when obtaining more precise individual adherence estimates, we can start characterising the determinants by integrating prescription datasets with rich individual-level data such as socioeconomic status, personality characteristics, and genetic profiles. The substantial year-to-year variability in adherence patterns in the current study indicates that medication-taking behaviour is shaped by both enduring patient characteristics and dynamic life circumstances. In future research, it is important to tease these effects apart.

Second, the strong effect of IMAS in our study points to the need for more personalised adherence interventions rather than population-level strategies. Our novel method using IMAS seems a promising approach to identify high-risk patients and to provide timely interventions, especially as IMAS had better predictive power compared to different demographic, health- and medication-related factors, even across medication classes. Therefore, we believe that IMAS can significantly contribute to more effective personalised risk models which take into account medication administration history across different health conditions and medications.

Generally, better medication adherence has been associated with a lower risk of several diagnoses^12,13^ and hospitalisation.^4,12,14^ We showed a 24% reduction in the risk of overall hospitalisation and a 27– 58% reduction in the incidence risk for several diseases among those with better adherence. However, the decrease in the incidence risk was not observed for all diseases. This highlights the nuanced relationship between adherence and health outcomes. Proper administration of necessary medications certainly helps to manage chronic disease, but patients with better adherence may visit their healthcare providers more frequently, resulting in earlier and potentially more diagnoses. At the same time, it could be hypothesized that a significant deterioration of health or an acute episode could have a positive effect on medication adherence. Decomposing such a complex causal network is an important next step in understanding the phenomenon. However, it requires a different set of methodologies and is out of the scope of the current paper.

The study has limitations that need to be acknowledged. One limitation of using prescription data is the lack of information of whether or how the medication was actually taken. Also, any dosing changes by doctors after the medication has already been purchased remain undetected. A crucial aspect of calculating adherence measures on prescription data is the accuracy and completeness of the records. In some countries, the completeness of prescription data is hindered by the diversity of insurers or pharmacies with independent non-linkable systems. However, that is not the case in our study, as the national prescription database stores information about all issued and dispensed prescription medications. Still, the day’s supply was missing for 38% of the prescriptions. The missing information was replaced with imputation potentially causing systematic bias. However, this potential medication-level systematic bias is taken into account in medication-based fixed effects and its influence on IMAS can be considered modest. Still, the imputation methods can be refined further to better reflect the dosing patterns for various ingredients, thus resulting in more accurate adherence measures.

At the same time, the strengths of our study are analysing more than 130 ingredients in a consistent manner in over 60 thousand patients, allowing us to detect more subtle effects and perform accurate between-ingredient comparisons. Moreover, our analysis takes into account a wide range of factors from medical records potentially influencing medication adherence, thus, reducing apparent confounding effects. The study provides a framework for further medication adherence research on real-world health databases modelling person-level variation while taking into account linked health events and other data modalities. Further aspects of medication adherence, such as primary adherence and persistence, should also be included in building more accurate prediction methods for identifying high-risk patients.

Our study highlights the value and possibility of integrating large-scale real-world data for proactive identification of at-risk patients. The study warrants a shift from specific medication and population-based focus towards a more patient-level perspective in adherence research. The predictiveness of the IMAS suggests the potential for personalised interventions in clinical practice and more efficient public health strategies and policies.

## Methods

### Data and setting

The dataset consisted of all electronic health records (EHR), healthcare service provision claims, and prescribed medications from 01.01.2012 to 31.12.2019 for randomly selected 10% of the Estonian population (N = 150 824). The EHR, claims and prescriptions data originated from three health databases with national coverage storing information from almost all healthcare settings (hospitals, specialists, family doctors, labs, pharmacies). EHR covers data from all private and state-owned healthcare providers regardless of the insurance of the patient, while claims covers only people with public health insurance (approximately 95% of the Estonian population^15^). The drug prescription database includes all prescribed drugs^16^ – their ingredient, Anatomical Therapeutic Chemical (ATC) code, product name and code, amount, administration guidelines, purchase date and location, a healthcare provider who issued the medication, and the International Statistical Classification of Diseases and Related Health Problems 10th Revision (ICD-10) code for the condition being treated. The databases were linked using a unique personal ID code given to all Estonian residents, and data was transferred to the Observational Medical Outcomes Partnership (OMOP) common data model (CDM) version 5.4.^17^ The overall workflow of the analysis is presented in Fig. 4.

**Fig. 4.**
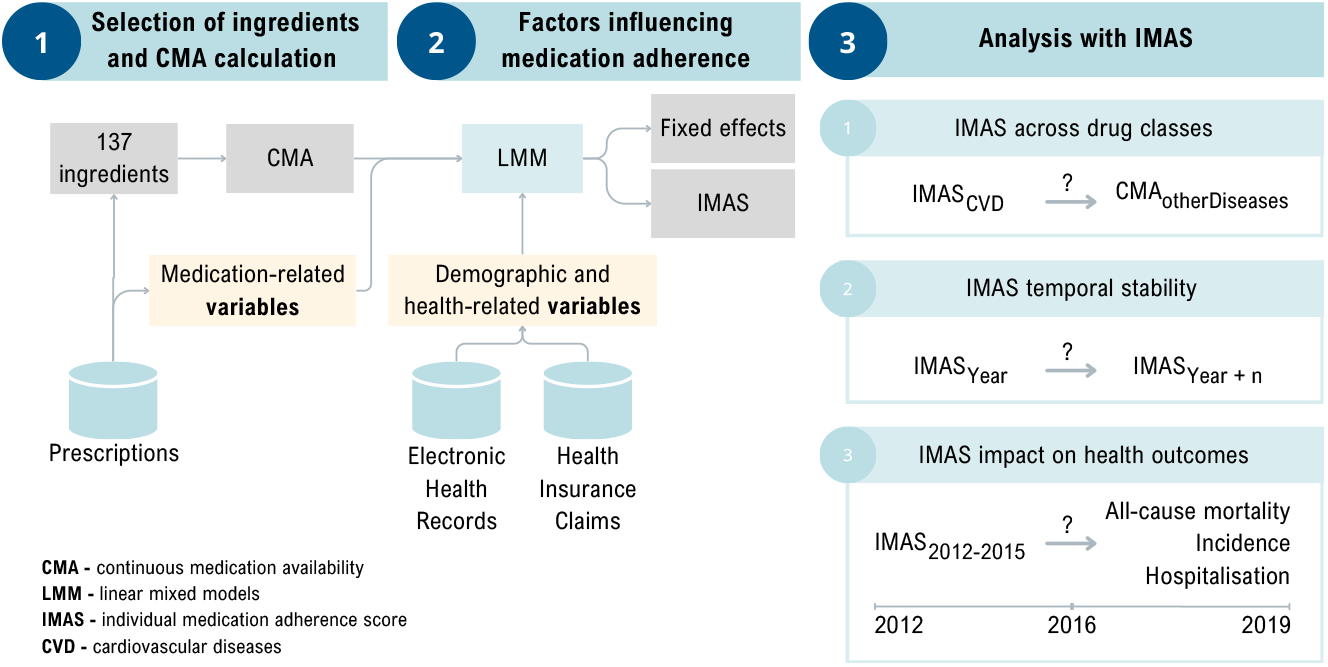
Workflow of the analysis.

### Calculation of medication adherence

First, 300 of the most prescribed ingredients or ingredient combinations were extracted from the database. These ingredients were evaluated independently by two pharmacists who classified them as 1) ingredients meant for long-term use and 2) others. The interobserver variability was assessed with intraclass correlation (r = 0.94, p < 0.05). In case of contradictions in the pharmacist assessment, the results were validated by a medical doctor. As a result, 137 ingredients for chronic conditions were identified and included in the adherence calculations (Fig. 4, step 1). As each prescription includes diagnosis info, the most frequent diagnosis for each ingredient was extracted. Based on medical expert opinion and previous studies,^18^ the ingredients were combined into ICD-10-based diagnosis groups (Supplementary Table 2).

For medication adherence calculations we applied continuous multiple interval measures of medication availability (CMA) on purchased prescriptions.^19^ Out of eight CMA measures, CMA5 was used as it considers the gap times in medication availability and assumes that the new refill is banked until the previous prescription is depleted.^19^ Moreover, it allows carry-over within the observation window, while any unused medication available at the end of the observation window is ignored in the calculations.^19^ The CMA5 value was calculated with a yearly interval from the beginning of the first purchase of the ingredient. At least one refill within 365 days was required to be included in the analysis. The days of supply for ingredients were calculated using the following information from the prescription: the amount of drugs in one package (e.g. number of tablets), the number of packages and the dosage prescribed by the doctor. This allowed us to calculate the number of days for which the patient had medication available. If treatment length information was present, the smaller from the calculated and provided treatment length values were used. If dosage was missing, then the mode of day’s supply value from existing data was used. The calculations were ingredient-based to minimise the potential effects of medication substitutions (e.g. original vs. generic drugs) and drug scheme changes. This means that if the person received one ingredient and the drug scheme was changed so that a combination of old and new ingredients was provided, then the CMA5 calculations continued for the old ingredient and in parallel started for the new ingredient. The CMA5 was calculated with AdhereR package.^20^

For descriptive statistics proportions, means with standard deviations, and the prevalence of good adherers (CMA ≥ 0.8) were calculated. The gender differences were analysed with the Mann-Whitney U test.

### Factors influencing medication adherence

To assess the effect of various demographic, health, and medication-related variables on CMA, we employed linear mixed models (LMM) using packages lme4 (v1.1-14)^21^ together with package rsq (v2.6)^22^ to calculate models’ R-square (Fig. 4, step 2). LMM was chosen for its ability to capture individual-level variability and its suitability for repeated measures. Subjects were included in the model as random effects, while all other variables were treated as fixed effects. This approach allowed us to account for both within-subject changes over time and between-subject differences. For LMM, the coefficient and 95% confidence intervals are reported.

The demographical fixed effects included in the model were gender (male/ female) and age categorised into five groups: 0–19 years, 20–39 years, 40–59 years, 60–79 years, and 80 years and above. Body mass index (BMI) was included in the model as a categorical variable with four categories: 1) under and normal weight (BMI < 24.9), 2) overweight (25.0 ≤ BMI < 29.9), 3) obese (BMI ≥ 30), and 4) unknown. The occurrence of hospitalisation (yes/ no), diagnosis of depression (yes/ no), number of all diagnoses, and number of different ingredients administered were assessed and included in the model on a yearly basis. The first occurrence of a comorbidity condition that belongs to the Charlson Comorbidity Index^23^ and the first occurrence of dementia or mental retardation diagnosis set the status for the following years. Dementia and mental retardation diagnoses were used to discriminate based on cognitive impairment, which has previously been identified as a risk factor for low medication adherence.^24^ Ingredient was included in the model as a categorical variable. Metoprolol was used as a reference category as this was the most frequently prescribed ingredient. For each ingredient, its administration route and consecutive year of ingredient administration were included in the model. As each ingredient was mainly prescribed for specific condition, a binary value for the most prevalent diagnosis was created. The combination of the ingredient and its most prevalent diagnoses was marked as 1, while all other combinations were 0. The interaction between 137 ingredients and the 12 most prevalent conditions was also included in the model. The Bonferroni correction was applied to the LMM to evaluate the statistical significance of the variables.

### Individual Medication Adherence Score (IMAS)

The Individual Medication Adherence Score (IMAS) is the person-specific effect extracted from LMM which included previously described demographic, health- and medication-related variables. IMAS represents an individual’s inherent medication-taking behaviour – their personal “baseline” adherence tendency that persists across prescriptions and time, after accounting for demographic, medication- and health-related factors.

### Analysis with IMAS

To determine whether IMAS calculated for ingredients related to one disease group helps to predict the medication adherence for other diseases, we first calculated LMM using only ingredients associated with different cardiovascular diseases (CVD) (hypertension (I10-I15), arrhythmias (I46-I49), heart failure (I50), ischemic heart disease (I20-I25), other diseases of the circulatory system (I67-I70)) (CVD model) (Fig. 4, step 3). Next, two LMMs were created to assess the effect on medication adherence: 1) LMM with all other ingredients and 2) LMM with all other ingredients and IMAS from the CVD model. Similarly to previous models demographical, health- and medication-related variables were added as fixed effects in all these models. This approach allowed us to evaluate how medication adherence calculated for some ingredients affects the medication adherence of other ingredients.

To assess the stability of the IMAS over time, we calculated IMAS using LMM with demographical, health- and medication-related variables as fixed effects for each year. Next, the correlation between yearly IMAS was calculated.

For survival analysis, we created LMM for medication adherence with demographical, health- and medication-related variables as fixed effects using data from 2012 to 2016 and calculated IMAS which was included in the Cox proportional hazards model to estimate incidence and hospitalisation for selected diseases (Supplementary Table 7) between 2017 and 2019. Only persons who were alive on 01.01.2017 and did not have an occurrence of chronic condition under observation or hospitalisation from 2012–2016 were included in the analysis. The diagnoses included in the analysis were based on previous studies^18,25^ and medical expert opinion. In the Cox proportional hazards model, the IMAS was continuous variable, and the model was controlled for year of birth and gender. The Bonferroni correction was applied to evaluate the significance of hazard ratios. The hazard ratios, together with 95% CI, are presented. Significant IMAS was visualised with Kaplan-Meier curve. All analyses were performed using R (v4.3.3).

The study was approved by the Research Ethics Committee of the University of Tartu (300/T-23) and the Estonian Committee on Bioethics and Human Research (1.1-12/653), and the requirement for informed consent was waived.

## Data Availability

There are legal restrictions on sharing de-identified data. According
to legislative regulation and data protection law in Estonia, the authors
cannot publicly release or share the data received from the health data registries in Estonia.

## Contributions

KM, RK, TT, HL, NU, SR, JV, JH, MO participated in the design of the study and methodology. The literature search was carried out by KM and HK. Data curators were ST, MO, MM. Software developments were done by JH, MO, MM. The analysis was conducted by KM, HL, RK. Figures were designed by MP. Funding acquisition – RK, SR, JV. The original draft was written by KM while all authors read and edited the manuscript and approved the final version.

## Acknowledgements

This study was co-funded by the European Union and Estonian Ministry of Education and Research via project TEM-TA72 and Estonian Research Council grants PRG1844 and PSG809.

## Data availability

The datasets generated and analysed during the current study are not publicly available due to legal restrictions on sharing de-identified data. According to legislative regulation and data protection law in Estonia, the authors cannot publicly release the data received from the health data registries in Estonia. However, the data can be requested by completing necessary applications in order to carry out research or an evaluation of public interest and acquiring the permission of the controller of the databases (https://www.tehik.ee/en/statistics). More information about data availability.

## Code availability

The underlying code for this study [and training/validation datasets] is not publicly available but may be made available to qualified researchers on reasonable request from the corresponding author.

