## Supplementary material for "Systematic evaluation of medication adherence determinants across 137 ingredients on population-level real-world health data"

**Supplementary Table 1.** Average CMA with 95% confidence intervals by ingredients

| Ingredient | Average CMA | 95% CI |
|---|---|---|
| agomelatine | 0.766 | 0.750…0.783 |
| albuterol | 0.423 | 0.414…0.432 |
| alendronate | 0.812 | 0.801…0.823 |
| alfuzosin | 0.733 | 0.720…0.747 |
| allopurinol | 0.727 | 0.720…0.734 |
| amiodarone | 0.781 | 0.770…0.793 |
| amlodipine | 0.796 | 0.793…0.799 |
| amylase | 0.453 | 0.436…0.469 |
| anastrozole | 0.901 | 0.883…0.918 |
| apixaban | 0.834 | 0.825…0.842 |
| aripiprazole | 0.794 | 0.766…0.822 |
| atenolol | 0.790 | 0.768…0.812 |
| atorvastatin | 0.787 | 0.782…0.792 |
| benserazide | 0.825 | 0.807…0.843 |
| betaxolol | 0.786 | 0.765…0.807 |
| bicalutamide | 0.820 | 0.803…0.838 |
| bimatoprost | 0.784 | 0.769…0.798 |
| bisoprolol | 0.768 | 0.756…0.780 |
| brinzolamide | 0.822 | 0.812…0.831 |
| bupropion | 0.797 | 0.781…0.813 |
| candesartan | 0.730 | 0.720…0.740 |
| carbamazepine | 0.719 | 0.704…0.733 |
| carvedilol | 0.776 | 0.757…0.795 |
| chlorprothixene | 0.829 | 0.804…0.853 |
| citalopram | 0.767 | 0.752…0.781 |
| clopidogrel | 0.865 | 0.858…0.872 |
| clozapine | 0.829 | 0.807…0.852 |
| cyclosporine | 0.866 | 0.836…0.897 |
| dabigatran | 0.763 | 0.746…0.780 |
| digoxin | 0.813 | 0.806…0.821 |
| donepezil | 0.805 | 0.781…0.828 |
| dorzolamide | 0.809 | 0.800…0.818 |
| doxazosin | 0.741 | 0.725…0.758 |
| duloxetine | 0.816 | 0.805…0.827 |
| dutasteride | 0.815 | 0.805…0.826 |
| empagliflozin | 0.828 | 0.813…0.843 |
| enalapril | 0.818 | 0.813…0.823 |
| escitalopram | 0.781 | 0.774…0.787 |
| febuxostat | 0.746 | 0.728…0.764 |
| felodipine | 0.845 | 0.833…0.857 |
| fluoxetine | 0.786 | 0.776…0.797 |
| flupenthixol | 0.690 | 0.665…0.714 |
| fluvastatin | 0.759 | 0.731…0.788 |
| formoterol | 0.633 | 0.624…0.642 |
| fosinopril | 0.826 | 0.818…0.833 |
| furosemide | 0.612 | 0.601…0.624 |
| gliclazide | 0.859 | 0.852…0.866 |
| glimepiride | 0.837 | 0.828…0.847 |

| | | |
|---|---|---|
| glycopyrronium | 0.755 | 0.734...0.776 |
| haloperidol | 0.633 | 0.614...0.651 |
| hydrochlorothiazide | 0.787 | 0.783...0.791 |
| hydroxychloroquine | 0.767 | 0.752...0.783 |
| indacaterol | 0.754 | 0.731...0.776 |
| indapamide | 0.796 | 0.791...0.801 |
| insulin aspart | 0.727 | 0.716...0.739 |
| insulin detemir | 0.715 | 0.701...0.728 |
| insulin glargine | 0.714 | 0.703...0.726 |
| insulin glulisine, human | 0.712 | 0.681...0.743 |
| insulin lispro | 0.764 | 0.744...0.783 |
| ipratropium | 0.505 | 0.486...0.524 |
| isosorbide | 0.767 | 0.758...0.777 |
| lacidipine | 0.799 | 0.789...0.810 |
| lamotrigine | 0.840 | 0.819...0.861 |
| latanoprost | 0.789 | 0.782...0.796 |
| leflunomide | 0.697 | 0.667...0.728 |
| lercanidipine | 0.836 | 0.828...0.845 |
| letrozole | 0.840 | 0.812...0.868 |
| levetiracetam | 0.899 | 0.875...0.923 |
| levodopa | 0.832 | 0.816...0.849 |
| levothyroxine | 0.883 | 0.879...0.887 |
| linagliptin | 0.844 | 0.832...0.856 |
| lipase | 0.453 | 0.436...0.469 |
| liraglutide | 0.693 | 0.672...0.715 |
| lisinopril | 0.775 | 0.759...0.790 |
| losartan | 0.822 | 0.814...0.830 |
| melperone hydrochloride | 0.724 | 0.708...0.741 |
| mesalamine | 0.699 | 0.672...0.725 |
| metformin | 0.795 | 0.791...0.799 |
| methimazole | 0.772 | 0.755...0.788 |
| methotrexate | 0.882 | 0.871...0.893 |
| methylphenidate | 0.652 | 0.630...0.675 |
| metoprolol | 0.788 | 0.785...0.791 |
| mirtazapine | 0.733 | 0.721...0.745 |
| montelukast | 0.588 | 0.575...0.602 |
| moxonidine | 0.607 | 0.589...0.626 |
| nafronyl | 0.698 | 0.682...0.714 |
| nebivolol | 0.749 | 0.744...0.754 |
| nifedipine | 0.725 | 0.705...0.745 |
| nitrendipine | 0.769 | 0.759...0.779 |
| nortriptyline | 0.696 | 0.670...0.721 |
| olanzapine | 0.803 | 0.786...0.820 |
| olmesartan | 0.786 | 0.778...0.793 |
| oxcarbazepine | 0.770 | 0.743...0.798 |
| paroxetine | 0.751 | 0.738...0.764 |
| pentoxifylline | 0.677 | 0.665...0.690 |
| perindopril | 0.804 | 0.800...0.808 |
| piracetam | 0.593 | 0.569...0.616 |
| pramipexole | 0.802 | 0.773...0.831 |

| | | |
|---|---|---|
| pravastatin | 0.770 | 0.741…0.799 |
| promazine | 0.824 | 0.801…0.848 |
| propafenone | 0.723 | 0.711…0.735 |
| propranolol | 0.717 | 0.699…0.735 |
| protease | 0.453 | 0.436…0.469 |
| ramipril | 0.800 | 0.796…0.804 |
| rasagiline | 0.857 | 0.835…0.878 |
| risperidone | 0.780 | 0.763…0.796 |
| rivaroxaban | 0.825 | 0.817…0.833 |
| rosuvastatin | 0.761 | 0.756…0.765 |
| saxagliptin | 0.839 | 0.809…0.868 |
| sertraline | 0.790 | 0.781…0.800 |
| simvastatin | 0.780 | 0.773…0.787 |
| sitagliptin | 0.822 | 0.811…0.833 |
| sotalol | 0.814 | 0.798…0.831 |
| spironolactone | 0.732 | 0.724…0.741 |
| sulfasalazine | 0.712 | 0.692…0.731 |
| tafluprost | 0.819 | 0.804…0.834 |
| tamoxifen | 0.874 | 0.854…0.894 |
| tamsulosin | 0.774 | 0.767…0.780 |
| telmisartan | 0.791 | 0.787…0.795 |
| theophylline | 0.668 | 0.649…0.687 |
| tianeptine | 0.671 | 0.658…0.684 |
| ticagrelor | 0.883 | 0.871…0.894 |
| timolol | 0.803 | 0.797…0.809 |
| tiotropium | 0.797 | 0.782…0.811 |
| torsemide | 0.646 | 0.638…0.653 |
| trandolapril | 0.807 | 0.787…0.827 |
| travoprost | 0.797 | 0.786…0.808 |
| trihexyphenidyl | 0.744 | 0.722…0.766 |
| trimetazidine dihydrochloride | 0.622 | 0.614…0.631 |
| ursodeoxycholate | 0.678 | 0.648…0.707 |
| valproate | 0.810 | 0.794…0.826 |
| valsartan | 0.805 | 0.795…0.815 |
| venlafaxine | 0.846 | 0.835…0.856 |
| verapamil | 0.831 | 0.820…0.841 |
| vildagliptin | 0.826 | 0.806…0.845 |
| vortioxetine | 0.805 | 0.783…0.827 |
| warfarin | 0.922 | 0.917…0.926 |

**Supplementary Table 2.** Ingredients included into the analysis, number of calculated continuous multiple interval measures of medication availability (CMA) measures, number of subjects, and most frequent diagnoses on the prescription

| Ingredient | N of CMAs | N subjects | Disease group | ICD-10 Chapter |
|---|---|---|---|---|
| anastrozole | 529 | 205 | Malignant neoplasm of breast (C50) | Neoplasms (C00-D48) |
| letrozole | 877 | 368 | | |
| tamoxifen | 747 | 324 | | |
| bicalutamide | 971 | 479 | Malignant neoplasm of prostate (C61) | |
| levothyroxine | 24153 | 5443 | Disorders of thyroid gland (E00-E07) | Endocrine, nutritional and metabolic diseases (E00-E90) |
| methimazole | 1161 | 688 | | |
| empagliflozin | 1342 | 672 | Diabetes (E10-E14) | |
| gliclazide | 8225 | 2150 | | |
| glimepiride | 5625 | 1459 | | |
| insulin aspart | 4764 | 1167 | | |
| insulin detemir | 3386 | 884 | | |
| insulin glargine | 4666 | 1195 | | |
| insulin glulisine, human | 756 | 179 | | |
| insulin lispro | 1621 | 374 | | |
| linagliptin | 2328 | 927 | | |
| liraglutide | 977 | 335 | | |
| metformin | 32311 | 7659 | | |
| metoprolol | 69748 | 18179 | | |
| saxagliptin | 445 | 170 | | |
| sitagliptin | 4208 | 1127 | | |
| vildagliptin | 893 | 349 | | |
| atorvastatin | 22550 | 7074 | Disorders of lipoprotein metabolism and other lipidaemias (E78) | |
| fluvastatin | 713 | 218 | | |
| pravastatin | 657 | 173 | | |
| rosuvastatin | 25418 | 7196 | | |
| simvastatin | 11751 | 3035 | | |
| donepezil | 759 | 335 | Dementia (F00-F03, G30, G31) | Mental and behavioural disorders (F00-F99) |
| melperone hydrochloride | 1764 | 835 | | |
| agomelatine | 1207 | 923 | Psychotic, mood and neurotic disorders (F20-F48, F90) | |
| aripiprazole | 640 | 235 | | |
| bupropion | 1268 | 745 | | |
| chlorprothixene | 943 | 307 | | |
| citalopram | 2204 | 926 | | |
| clozapine | 1042 | 269 | | |
| duloxetine | 2842 | 1368 | | |
| escitalopram | 8264 | 4176 | | |
| fluoxetine | 3496 | 1745 | | |
| flupenthixol | 864 | 418 | | |
| haloperidol | 2153 | 1003 | | |
| methylphenidate | 939 | 420 | | |
| mirtazapine | 3834 | 1900 | | |
| nortriptyline | 810 | 336 | | |
| olanzapine | 1729 | 634 | | |

| | | | | |
|---|---|---|---|---|
| paroxetine | 2774 | 1140 | | |
| promazine | 847 | 327 | | |
| risperidone | 1740 | 727 | | |
| sertraline | 4203 | 2238 | | |
| tianeptine | 3025 | 1861 | | |
| trihexyphenidyl | 1435 | 455 | | |
| venlafaxine | 2923 | 1326 | | |
| vortioxetine | 548 | 414 | | |
| benserazide | 1635 | 503 | Parkinson disease (G20) | Diseases of the nervous system (G00-G99) |
| levodopa | 2055 | 579 | | |
| pramipexole | 769 | 276 | | |
| rasagiline | 804 | 239 | | |
| carbamazepine | 3925 | 1247 | Epilepsy (G40-G42) | |
| lamotrigine | 1047 | 350 | | |
| levetiracetam | 454 | 145 | | |
| oxcarbazepine | 961 | 279 | | |
| valproate | 2526 | 721 | | |
| betaxolol | 1270 | 441 | Glaucoma (H40-H42) | Diseases of the eye and adnexa (H00-H59) |
| bimatoprost | 2120 | 733 | | |
| brinzolamide | 4839 | 1524 | | |
| dorzolamide | 5701 | 1788 | | |
| latanoprost | 8627 | 2614 | | |
| tafluprost | 1705 | 666 | | |
| timolol | 15153 | 4024 | | |
| travoprost | 3966 | 1170 | | |
| amlodipine | 53205 | 14911 | Hypertension (I10-I15) | Diseases of the circulatory system (I00-I99) |
| atenolol | 1500 | 388 | | |
| bisoprolol | 3370 | 1341 | | |
| candesartan | 6500 | 1946 | | |
| enalapril | 20166 | 5430 | | |
| felodipine | 3446 | 920 | | |
| fosinopril | 9737 | 2553 | | |
| hydrochlorothiazide | 41332 | 10915 | | |
| indapamide | 24722 | 8396 | | |
| lacidipine | 4852 | 1367 | | |
| lercanidipine | 5478 | 1812 | | |
| lisinopril | 2326 | 721 | | |
| losartan | 7254 | 1911 | | |
| moxonidine | 2564 | 1113 | | |
| nebivolol | 21924 | 6831 | | |
| nifedipine | 1912 | 822 | | |
| nitrendipine | 6121 | 2074 | | |
| olmesartan | 9696 | 3305 | | |
| perindopril | 29351 | 10107 | | |
| propranolol | 1705 | 798 | | |
| ramipril | 32550 | 10038 | | |
| telmisartan | 34302 | 9070 | | |
| trandolapril | 1172 | 387 | | |

| | | | | |
|---|---|---|---|---|
| valsartan | 5482 | 1586 | | |
| verapamil | 4085 | 1316 | | |
| isosorbide | 6460 | 2078 | Ischemic heart disease (I20-I25) | |
| trimetazidine dihydrochloride | 9032 | 3522 | | |
| amiodarone | 3275 | 1346 | Arrhythmias (I46-I49) | |
| apixaban | 3471 | 1929 | | |
| dabigatran | 2026 | 956 | | |
| propafenone | 4708 | 1599 | | |
| rivaroxaban | 6706 | 2977 | | |
| sotalol | 2186 | 660 | | |
| warfarin | 10576 | 2976 | | |
| carvedilol | 1548 | 500 | Heart failure (I50) | |
| digoxin | 7334 | 2453 | | |
| furosemide | 4623 | 2321 | | |
| spironolactone | 7352 | 3241 | | |
| torsemide | 11610 | 5492 | | |
| nafronyl | 2245 | 1010 | Other diseases of the circulatory system (I67-I70) | |
| pentoxifylline | 3834 | 1612 | | |
| piracetam | 965 | 591 | | |
| albuterol | 9250 | 5330 | Chronic diseases of respiratory system (J43-J47) | Diseases of the respiratory system (J00-J99) |
| formoterol | 8446 | 2989 | | |
| glycopyrronium | 1038 | 431 | | |
| indacaterol | 911 | 389 | | |
| ipratropium | 1766 | 776 | | |
| montelukast | 3232 | 1705 | | |
| theophylline | 1810 | 752 | | |
| tiotropium | 2012 | 812 | | |
| amylase | 2384 | 1207 | Diseases of the digestive system (K00-K93) | Diseases of the digestive system (K00-K93) |
| lipase | 2384 | 1207 | | |
| mesalamine | 944 | 306 | | |
| protease | 2384 | 1207 | | |
| ursodeoxycholate | 667 | 343 | | |
| hydroxychloroquine | 1869 | 704 | Rheumatoid arthritis and related disorders (M05, M06, M08, M13, M30-35, M45) | Diseases of the musculoskeletal system and connective tissue (M00-M99) |
| leflunomide | 731 | 239 | | |
| methotrexate | 2803 | 1081 | | |
| sulfasalazine | 1536 | 552 | | |
| allopurinol | 11183 | 4164 | Gout (M10) | |
| febuxostat | 1277 | 745 | | |
| alendronate | 3890 | 1366 | Osteoporosis (M80, M81) | |
| alfuzosin | 2180 | 1214 | Diseases of male genital organs (N40-N51) | Diseases of the genitourinary system (N00-N99) |
| doxazosin | 2220 | 781 | | |
| dutasteride | 3836 | 1337 | | |
| tamsulosin | 11020 | 3920 | | |
| clopidogrel | 4927 | 2428 | Presence of cardiac and vascular implants and grafts (Z95) | Factors influencing health status and contact with health services (Z00-Z99) |
| cyclosporine | 444 | 153 | | |
| ticagrelor | 1038 | 796 | | |

**Supplementary Table 3.** Average age by ingredient

| Ingredient | Average age | 95% CI |
|---|---|---|
| agomelatine | 47.1 | 46.0...48.1 |
| albuterol | 30.7 | 29.9...31.5 |
| alendronate | 72.4 | 71.8...72.9 |
| alfuzosin | 55.5 | 54.5...56.5 |
| allopurinol | 67.2 | 66.8...67.6 |
| amiodarone | 71.4 | 70.8...72.1 |
| amlodipine | 66.8 | 66.6...67.0 |
| amylase | 63.8 | 62.8...64.8 |
| anastrozole | 68.5 | 66.9...70.1 |
| apixaban | 75.7 | 75.2...76.2 |
| aripiprazole | 35.9 | 33.9...37.8 |
| atenolol | 68.4 | 67.1...69.7 |
| atorvastatin | 67.7 | 67.4...68.0 |
| benserazide | 76.3 | 75.5...77.1 |
| betaxolol | 72.0 | 70.8...73.2 |
| bicalutamide | 74.5 | 73.8...75.3 |
| bimatoprost | 71.8 | 71.0...72.7 |
| bisoprolol | 67.4 | 66.7...68.2 |
| brinzolamide | 73.2 | 72.7...73.8 |
| bupropion | 40.5 | 39.4...41.6 |
| candesartan | 63.1 | 62.5...63.7 |
| carbamazepine | 57.3 | 56.2...58.4 |
| carvedilol | 68.9 | 67.7...70.0 |
| chlorprothixene | 57.5 | 55.6...59.3 |
| citalopram | 54.9 | 53.8...56.1 |
| clopidogrel | 71.0 | 70.5...71.4 |
| clozapine | 53.5 | 51.4...55.5 |
| cyclosporine | 49.7 | 46.8...52.6 |
| dabigatran | 73.0 | 72.2...73.7 |
| digoxin | 77.1 | 76.7...77.5 |
| donepezil | 77.4 | 76.5...78.3 |
| dorzolamide | 73.7 | 73.1...74.3 |
| doxazosin | 68.8 | 67.8...69.7 |
| duloxetine | 51.0 | 50.1...51.9 |
| dutasteride | 71.7 | 71.2...72.2 |
| empagliflozin | 62.4 | 61.7...63.2 |
| enalapril | 70.1 | 69.8...70.5 |
| escitalopram | 45.7 | 45.2...46.2 |
| febuxostat | 66.7 | 65.7...67.7 |
| felodipine | 72.0 | 71.2...72.8 |
| fluoxetine | 47.1 | 46.2...47.9 |
| flupenthixol | 56.2 | 54.5...58.0 |
| fluvastatin | 65.6 | 64.0...67.2 |
| formoterol | 54.9 | 54.1...55.6 |
| fosinopril | 68.8 | 68.3...69.3 |
| furosemide | 74.0 | 73.4...74.6 |
| gliclazide | 69.3 | 68.7...69.8 |
| glimepiride | 67.5 | 66.9...68.0 |
| glycopyrronium | 68.1 | 67.1...69.1 |
| haloperidol | 69.3 | 68.1...70.4 |

| | | |
|---|---|---|
| hydrochlorothiazide | 68.4 | 68.2...68.7 |
| hydroxychloroquine | 50.6 | 49.1...52.1 |
| indacaterol | 68.6 | 67.6...69.7 |
| indapamide | 65.2 | 64.9...65.4 |
| insulin aspart | 58.8 | 57.6...60.0 |
| insulin detemir | 58.5 | 57.3...59.7 |
| insulin glargine | 60.5 | 59.6...61.5 |
| insulin glulisine, human | 48.3 | 45.0...51.7 |
| insulin lispro | 57.5 | 55.4...59.6 |
| ipratropium | 67.3 | 66.3...68.3 |
| isosorbide | 77.3 | 76.9...77.7 |
| lacidipine | 71.6 | 70.9...72.2 |
| lamotrigine | 39.9 | 37.9...41.9 |
| latanoprost | 72.8 | 72.4...73.2 |
| leflunomide | 59.5 | 57.9...61.1 |
| lercanidipine | 69.4 | 68.8...69.9 |
| letrozole | 65.0 | 63.5...66.6 |
| levetiracetam | 31.7 | 28.1...35.2 |
| levodopa | 76.2 | 75.4...77.0 |
| levothyroxine | 59.3 | 58.8...59.8 |
| linagliptin | 69.3 | 68.5...70.0 |
| lipase | 63.8 | 62.8...64.8 |
| liraglutide | 60.7 | 59.6...61.8 |
| lisinopril | 65.1 | 64.2...66.0 |
| losartan | 67.6 | 67.1...68.2 |
| melperone hydrochloride | 73.0 | 71.8...74.2 |
| mesalamine | 46.1 | 43.9...48.2 |
| metformin | 65.6 | 65.3...65.9 |
| methimazole | 58.2 | 56.9...59.6 |
| methotrexate | 54.4 | 53.2...55.6 |
| methylphenidate | 13.6 | 12.9...14.3 |
| metoprolol | 68.7 | 68.5...68.9 |
| mirtazapine | 52.7 | 51.9...53.6 |
| montelukast | 23.4 | 22.2...24.5 |
| moxonidine | 69.0 | 68.2...69.7 |
| nafronyl | 72.2 | 71.4...72.9 |
| nebivolol | 59.5 | 59.1...59.8 |
| nifedipine | 74.8 | 74.0...75.6 |
| nitrendipine | 74.7 | 74.2...75.2 |
| nortriptyline | 60.5 | 58.3...62.7 |
| olanzapine | 48.7 | 47.3...50.1 |
| olmesartan | 64.8 | 64.4...65.2 |
| oxcarbazepine | 42.7 | 40.0...45.5 |
| paroxetine | 49.4 | 48.4...50.4 |
| pentoxifylline | 71.3 | 70.6...72.0 |
| perindopril | 64.3 | 64.0...64.5 |
| piracetam | 64.0 | 62.4...65.6 |
| pramipexole | 64.8 | 63.0...66.7 |
| pravastatin | 73.9 | 72.3...75.5 |
| promazine | 57.5 | 55.7...59.4 |
| propafenone | 69.3 | 68.8...69.9 |
| propranolol | 54.7 | 53.3...56.0 |
| protease | 63.8 | 62.8...64.8 |

| | | |
|---|---|---|
| ramipril | 68.4 | 68.2...68.7 |
| rasagiline | 74.0 | 72.8...75.2 |
| risperidone | 55.5 | 53.6...57.5 |
| rivaroxaban | 72.4 | 71.9...72.8 |
| rosuvastatin | 65.8 | 65.5...66.1 |
| saxagliptin | 63.1 | 61.4...64.8 |
| sertraline | 42.9 | 42.1...43.8 |
| simvastatin | 70.4 | 70.0...70.8 |
| sitagliptin | 65.7 | 65.0...66.3 |
| sotalol | 71.1 | 70.2...72.1 |
| spironolactone | 73.5 | 73.1...74.0 |
| sulfasalazine | 54.2 | 52.9...55.5 |
| tafluprost | 69.2 | 68.3...70.2 |
| tamoxifen | 56.1 | 54.3...57.9 |
| tamsulosin | 69.2 | 68.8...69.6 |
| telmisartan | 64.7 | 64.5...65.0 |
| theophylline | 74.0 | 73.2...74.9 |
| tianeptine | 53.6 | 52.8...54.5 |
| ticagrelor | 65.6 | 64.8...66.4 |
| timolol | 72.0 | 71.6...72.4 |
| tiotropium | 65.2 | 64.4...66.0 |
| torsemide | 75.5 | 75.2...75.8 |
| trandolapril | 63.9 | 62.6...65.2 |
| travoprost | 73.1 | 72.5...73.8 |
| trihexyphenidyl | 58.3 | 56.7...59.9 |
| trimetazidine dihydrochloride | 69.0 | 68.6...69.5 |
| ursodeoxycholate | 55.3 | 53.3...57.3 |
| valproate | 40.4 | 38.8...42.0 |
| valsartan | 65.5 | 64.8...66.1 |
| venlafaxine | 45.0 | 44.1...45.9 |
| verapamil | 69.2 | 68.5...69.9 |
| vildagliptin | 62.8 | 61.6...64.0 |
| vortioxetine | 43.0 | 41.6...44.4 |
| warfarin | 71.1 | 70.7...71.5 |

**Supplementary Table 4.** Estimates of demographic, health- and medication-related variables on CMA in linear mixed model

| Variable | Estimate | SE | 95% CI | P-value | Sig[1] |
|---|---|---|---|---|---|
| Intercept | 0.660 | 0.004 | 0.652...0.668 | 0.000 | * |
| Gender (Ref: Female) | | | | | |
| Male | -0.001 | 0.001 | -0.004...0.001 | 0.229 | |
| Age (Ref: 0-19 years) | | | | | |
| 20-39 years | 0.081 | 0.003 | 0.075...0.087 | 0.000 | * |
| 40-59 years | 0.079 | 0.003 | 0.073...0.085 | 0.000 | * |
| 60-79 years | 0.098 | 0.003 | 0.091...0.104 | 0.000 | * |
| 80+ years | 0.107 | 0.003 | 0.100...0.113 | 0.000 | * |
| Body mass index (Ref: obese) | | | | | |
| BMI: Overweight | -0.010 | 0.002 | -0.014...-0.006 | 0.000 | * |
| BMI: Under or normal weight | -0.009 | 0.002 | -0.014...-0.005 | 0.000 | * |
| BMI: Unknown | -0.011 | 0.002 | -0.014...-0.008 | 0.000 | * |
| Year of administation (Ref: year 1) | | | | | |
| Year 2 | 0.043 | 0.001 | 0.041...0.044 | 0.000 | * |
| Year 3 | 0.056 | 0.001 | 0.055...0.058 | 0.000 | * |
| Year 4 | 0.064 | 0.001 | 0.062...0.065 | 0.000 | * |
| Year 5 | 0.067 | 0.001 | 0.065...0.069 | 0.000 | * |
| Year 6 | 0.070 | 0.001 | 0.068...0.072 | 0.000 | * |
| Year 7 | 0.076 | 0.001 | 0.074...0.078 | 0.000 | * |
| Hospitalisation (Ref: No) | 0.009 | 0.001 | 0.007...0.010 | 0.000 | * |
| Depression (Ref: No) | 0.005 | 0.001 | 0003...0.007 | 0.000 | * |
| Dementia or retardation (Ref: No) | 0.004 | 0.002 | 0.000...0.009 | 0.053 | |
| Comorbidity (Ref: No) | 0.000 | 0.001 | -0.002...0.002 | 0.851 | |
| Number of chronic ingredients | 0.001 | 0.000 | 0.001...0.002 | 0.000 | * |
| Number of diagnoses | 0.000 | 0.000 | 0.000...0.000 | 0.004 | |
| Administration route | | | | | |
| Inhalation | 0.137 | 0.006 | 0.124...0.149 | 0.000 | * |
| Intramuscular | -0.293 | 0.013 | -0.319...-0.268 | 0.000 | * |
| Intravenous | -0.268 | 0.140 | -0.542...0.006 | 0.055 | |
| Mixed | 0.060 | 0.007 | 0.047...0.073 | 0.000 | * |
| Ocular | 0.026 | 0.014 | -0.001...0.053 | 0.062 | |
| Rectal | -0.213 | 0.031 | -0.274...-0.152 | 0.000 | * |
| Subcutaneous | 0.011 | 0.058 | -0.104...0.125 | 0.857 | |
| Ingredient (Ref: Metoprolol) | | | | | |
| Agomelatine | 0.021 | 0.008 | 0.005...0.038 | 0.010 | |
| Albuterol | -0.393 | 0.006 | -0.405...-0.382 | 0.000 | * |
| Alendronate | -0.048 | 0.019 | -0.085...-0.010 | 0.012 | |
| Alfuzosin | -0.014 | 0.011 | -0.036...0.008 | 0.210 | |
| Allopurinol | -0.048 | 0.006 | -0.060...-0.036 | 0.000 | * |
| Amiodarone | -0.024 | 0.008 | -0.040...-0.008 | 0.004 | |
| Amlodipine | 0.010 | 0.004 | 0.001...0.018 | 0.030 | |
| Amylase | -0.274 | 0.006 | -0.286...-0.263 | 0.000 | * |
| Anastrozole | -0.183 | 0.021 | -0.224...-0.142 | 0.000 | * |
| Apixaban | 0.027 | 0.011 | 0.005...0.050 | 0.017 | |
| Aripiprazole | -0.011 | 0.010 | -0.031...0.008 | 0.251 | |
| Atenolol | -0.026 | 0.017 | -0.060...0.007 | 0.121 | |
| Atorvastatin | 0.011 | 0.003 | 0.005...0.017 | 0.000 | * |
| Benserazide | -0.048 | 0.016 | -0.079...-0.017 | 0.003 | |
| Betaxolol | -0.187 | 0.075 | -0.333...-0.041 | 0.012 | |

| | | | | | |
|---|---|---|---|---|---|
| Bicalutamide | 0.001 | 0.117 | -0.228...0.231 | 0.990 | |
| Bimatoprost | 0.045 | 0.218 | -0.383...0.472 | 0.838 | |
| Bisoprolol | 0.000 | 0.006 | -0.012...0.012 | 0.982 | |
| Brinzolamide | -0.356 | 0.089 | -0.531...-0.181 | 0.000 | * |
| Bupropion | 0.032 | 0.009 | 0.014...0.050 | 0.000 | * |
| Candesartan | -0.075 | 0.012 | -0.098...-0.053 | 0.000 | * |
| Carbamazepine | -0.106 | 0.006 | -0.118...-0.094 | 0.000 | * |
| Carvedilol | -0.003 | 0.011 | -0.024...0.019 | 0.817 | |
| Chlorprothixene | 0.033 | 0.008 | 0.018...0.048 | 0.000 | * |
| Citalopram | -0.021 | 0.006 | -0.033...-0.008 | 0.001 | |
| Clopidogrel | 0.042 | 0.004 | 0.034...0.050 | 0.000 | * |
| Clozapine | -0.012 | 0.008 | -0.028...0.004 | 0.133 | |
| Cyclosporine | 0.037 | 0.016 | 0.005...0.069 | 0.023 | |
| Dabigatran | -0.067 | 0.010 | -0.087...-0.046 | 0.000 | * |
| Digoxin | 0.012 | 0.010 | -0.008...0.031 | 0.233 | |
| Donepezil | 0.024 | 0.012 | 0.001...0.046 | 0.043 | |
| Dorzolamide | -0.081 | 0.041 | -0.162...-0.001 | 0.047 | |
| Doxazosin | -0.041 | 0.009 | -0.059...-0.022 | 0.000 | * |
| Duloxetine | 0.039 | 0.005 | 0.029...0.049 | 0.000 | * |
| Dutasteride | 0.016 | 0.014 | -0.012...0.044 | 0.267 | |
| Empagliflozin | -0.153 | 0.076 | -0.301...-0.005 | 0.043 | |
| Enalapril | 0.019 | 0.007 | 0.005...0.032 | 0.007 | |
| Escitalopram | 0.009 | 0.004 | 0.001...0.017 | 0.027 | |
| Febuxostat | 0.003 | 0.017 | -0.031...0.036 | 0.878 | |
| Felodipine | 0.012 | 0.020 | -0.026...0.050 | 0.542 | |
| Fluoxetine | 0.007 | 0.005 | -0.004...0.017 | 0.208 | |
| Flupenthixol | -0.073 | 0.008 | -0.090...-0.057 | 0.000 | * |
| Fluvastatin | -0.031 | 0.013 | -0.055...-0.006 | 0.016 | |
| Formoterol | -0.237 | 0.009 | -0.255...-0.218 | 0.000 | * |
| Fosinopril | 0.010 | 0.009 | -0.008...0.027 | 0.285 | |
| Furosemide | -0.151 | 0.008 | -0.166...-0.136 | 0.000 | * |
| Gliclazide | 0.046 | 0.024 | -0.001...0.092 | 0.053 | |
| Glimepiride | 0.032 | 0.037 | -0.042...0.105 | 0.395 | |
| Glycopyrronium | -0.189 | 0.012 | -0.213...-0.165 | 0.000 | * |
| Haloperidol | -0.131 | 0.005 | -0.142...-0.121 | 0.000 | * |
| Hydrochlorothiazide | -0.023 | 0.005 | -0.032...-0.013 | 0.000 | * |
| Hydroxychloroquine | -0.004 | 0.007 | -0.017...0.010 | 0.595 | |
| Indacaterol | -0.195 | 0.013 | -0.220...-0.170 | 0.000 | * |
| Indapamide | 0.018 | 0.006 | 0.007...0.029 | 0.002 | |
| Insulin aspart | -0.060 | 0.059 | -0.176...0.056 | 0.310 | |
| Insulin detemir | -0.091 | 0.059 | -0.207...0.025 | 0.123 | |
| Insulin glargine | -0.102 | 0.059 | -0.218...0.014 | 0.084 | |
| Insulin glulisine, human | -0.060 | 0.060 | -0.177...0.058 | 0.319 | |
| Insulin lispro | -0.081 | 0.059 | -0.198...0.035 | 0.172 | |
| Ipratropium | -0.443 | 0.012 | -0.467...-0.419 | 0.000 | * |
| Isosorbide | -0.031 | 0.005 | -0.041...-0.020 | 0.000 | * |
| Lacidipine | 0.013 | 0.017 | -0.021...0.046 | 0.456 | |
| Lamotrigine | 0.004 | 0.008 | -0.013...0.020 | 0.642 | |
| Latanoprost | -0.059 | 0.054 | -0.165...0.047 | 0.277 | |
| Leflunomide | -0.090 | 0.009 | -0.109...-0.072 | 0.000 | * |
| Lercanidipine | 0.049 | 0.011 | 0.026...0.071 | 0.000 | * |
| Letrozole | -0.193 | 0.019 | -0.231...-0.155 | 0.000 | * |
| Levetiracetam | 0.012 | 0.012 | -0.012...0.036 | 0.325 | |

| | | | | | |
|---|---|---|---|---|---|
| Levodopa | -0.046 | 0.016 | -0.077...-0.015 | 0.004 | |
| Levothyroxine | 0.116 | 0.004 | 0.107...0.124 | 0.000 | * |
| Linagliptin | -0.073 | 0.063 | -0.195...0.050 | 0.246 | |
| Lipase | -0.274 | 0.006 | -0.286...-0.263 | 0.000 | * |
| Lisinopril | 0.005 | 0.024 | -0.043...0.052 | 0.840 | |
| Losartan | 0.034 | 0.012 | 0.009...0.058 | 0.007 | |
| Melperone hydrochloride | -0.055 | 0.006 | -0.067...-0.042 | 0.000 | * |
| Mesalamine | -0.104 | 0.016 | -0.135...-0.073 | 0.000 | * |
| Metformin | -0.035 | 0.007 | -0.048...-0.022 | 0.000 | * |
| Methimazole | -0.026 | 0.027 | -0.078...0.027 | 0.343 | |
| Methotrexate | 0.098 | 0.007 | 0.085...0.111 | 0.000 | * |
| Methylphenidate | 0.008 | 0.071 | -0.131...0.148 | 0.906 | |
| Mirtazapine | -0.044 | 0.005 | -0.054...-0.035 | 0.000 | * |
| Montelukast | -0.141 | 0.008 | -0.157...-0.125 | 0.000 | * |
| Moxonidine | -0.079 | 0.010 | -0.098...-0.060 | 0.000 | * |
| Nafronyl | -0.073 | 0.007 | -0.085...-0.060 | 0.000 | * |
| Nebivolol | -0.010 | 0.006 | -0.022...0.001 | 0.070 | |
| Nifedipine | 0.014 | 0.017 | -0.019...0.048 | 0.403 | |
| Nitrendipine | -0.021 | 0.011 | -0.042...-0.001 | 0.044 | |
| Nortriptyline | -0.092 | 0.009 | -0.110...-0.075 | 0.000 | * |
| Olanzapine | 0.008 | 0.006 | -0.004...0.021 | 0.183 | |
| Olmesartan | 0.017 | 0.010 | -0.002...0.036 | 0.072 | |
| Oxcarbazepine | -0.069 | 0.009 | -0.087...-0.050 | 0.000 | * |
| Paroxetine | -0.027 | 0.006 | -0.038...-0.016 | 0.000 | * |
| Pentoxifylline | -0.102 | 0.006 | -0.114...-0.091 | 0.000 | * |
| Perindopril | 0.024 | 0.006 | 0.013...0.035 | 0.000 | * |
| Piracetam | -0.122 | 0.008 | -0.137...-0.106 | 0.000 | * |
| Pramipexole | -0.002 | 0.013 | -0.027...0.023 | 0.873 | |
| Pravastatin | -0.041 | 0.017 | -0.074...-0.008 | 0.016 | |
| Promazine | 0.022 | 0.008 | 0.006...0.037 | 0.006 | |
| Propafenone | -0.060 | 0.007 | -0.074...-0.045 | 0.000 | * |
| Propranolol | -0.040 | 0.007 | -0.054...-0.025 | 0.000 | * |
| Protease | -0.274 | 0.006 | -0.286...-0.263 | 0.000 | * |
| Ramipril | 0.013 | 0.004 | 0.005...0.021 | 0.001 | |
| Rasagiline | -0.022 | 0.017 | -0.055...0.011 | 0.184 | |
| Risperidone | -0.021 | 0.006 | -0.033...-0.009 | 0.001 | |
| Rivaroxaban | 0.026 | 0.006 | 0.015...0.037 | 0.000 | * |
| Rosuvastatin | -0.007 | 0.003 | -0.013...0.000 | 0.036 | |
| Saxagliptin | -0.077 | 0.072 | -0.218...0.065 | 0.289 | |
| Sertraline | 0.023 | 0.005 | 0.013...0.032 | 0.000 | * |
| Simvastatin | -0.011 | 0.004 | -0.019...-0.004 | 0.004 | |
| Sitagliptin | -0.083 | 0.042 | -0.166...0.000 | 0.050 | |
| Sotalol | 0.029 | 0.008 | 0.013...0.045 | 0.000 | * |
| Spironolactone | -0.054 | 0.005 | -0.064...-0.043 | 0.000 | * |
| Sulfasalazine | -0.070 | 0.008 | -0.085...-0.054 | 0.000 | * |
| Tafluprost | -0.338 | 0.072 | -0.480...-0.196 | 0.000 | * |
| Tamoxifen | -0.143 | 0.018 | -0.178...-0.108 | 0.000 | * |
| Tamsulosin | -0.008 | 0.006 | -0.020...0.004 | 0.184 | |
| Telmisartan | 0.013 | 0.005 | 0.002...0.023 | 0.018 | |
| Theophylline | -0.152 | 0.009 | -0.170...-0.134 | 0.000 | * |
| Tianeptine | -0.094 | 0.005 | -0.104...-0.083 | 0.000 | * |
| Ticagrelor | 0.028 | 0.008 | 0.011...0.044 | 0.001 | |
| Timolol | -0.138 | 0.035 | -0.206...-0.069 | 0.000 | * |

| | | | | | |
|---|---|---|---|---|---|
| Tiotropium | -0.162 | 0.011 | -0.185...-0.140 | 0.000 | * |
| Torsemide | -0.138 | 0.004 | -0.146...-0.129 | 0.000 | * |
| Trandolapril | 0.034 | 0.025 | -0.014...0.083 | 0.167 | |
| Travoprost | -0.371 | 0.072 | -0.512...-0.229 | 0.000 | * |
| Trihexyphenidyl | -0.056 | 0.007 | -0.069...-0.043 | 0.000 | * |
| Trimetazidine dihydrochloride | -0.127 | 0.004 | -0.135...-0.118 | 0.000 | * |
| Ursodeoxycholate | -0.138 | 0.014 | -0.165...-0.111 | 0.000 | * |
| Valproate | -0.026 | 0.006 | -0.038...-0.013 | 0.000 | * |
| Valsartan | 0.021 | 0.013 | -0.003...0.046 | 0.089 | |
| Venlafaxine | 0.064 | 0.006 | 0.053...0.075 | 0.000 | * |
| Verapamil | 0.037 | 0.008 | 0.021...0.053 | 0.000 | * |
| Vildagliptin | -0.161 | 0.101 | -0.359...0.036 | 0.110 | |
| Vortioxetine | 0.038 | 0.011 | 0.016...0.060 | 0.001 | |
| Warfarin | 0.120 | 0.004 | 0.112...0.129 | 0.000 | * |
| Disease (Ref: No) | | | | | |
| Malignant neoplasm of breast (C50) | 0.300 | 0.019 | 0.263...0.337 | 0.000 | * |
| Malignant neoplasm of prostate (C61) | 0.022 | 0.117 | -0.208...0.252 | 0.853 | |
| Other hypothyroidism (E03) | -0.016 | 0.004 | -0.024...-0.008 | 0.000 | * |
| Hyperthyroidism (E05) | 0.021 | 0.028 | -0.033...0.075 | 0.445 | |
| Type 1 diabetes mellitus (E10) | 0.005 | 0.009 | -0.013...0.022 | 0.596 | |
| Type 2 diabetes mellitus (E11) | 0v203 | 0.101 | 0.005...0.401 | 0.044 | |
| Disorders of lipoprotein metabolism and other lipidaemias (E78) | -0.004 | 0.004 | -0.012...0.004 | 0.347 | |
| Dementia in Alzheimer disease (F00) | -0.004 | 0.017 | -0.037...0.030 | 0.837 | |
| Vascular dementia (F01) | 0.001 | 0.010 | -0.020...0.021 | 0.958 | |
| Other mental disorders due to brain damage and dysfunction and to physical disease (F06) | 0.009 | 0.011 | -0.013...0.031 | 0.435 | |
| Schizophrenia (F20) | 0.029 | 0.005 | 0.019...0.040 | 0.000 | * |
| Persistent delusional disorders (F22) | 0.031 | 0.019 | -0.007...0.069 | 0.109 | |
| Depressive episode (F32) | -0.003 | 0.020 | -0.041...0.035 | 0.881 | |
| Recurrent depressive disorder (F33) | 0.002 | 0.003 | -0.005...0.008 | 0.641 | |
| Other anxiety disorders (F41) | 0.015 | 0.003 | 0.009...0.021 | 0.000 | * |
| Hyperkinetic disorders (F90) | -0.048 | 0.072 | -0.189...0.093 | 0.503 | |
| Parkinson disease (G20) | 0.087 | 0.015 | 0.057...0.117 | 0.000 | * |
| Other extrapyramidal and movement disorders (G25) | -0.003 | 0.012 | -0.026...0.021 | 0.830 | |
| Alzheimer disease (G30) | 0.004 | 0.021 | -0.037...0.044 | 0.853 | |
| Epilepsy (G40) | 0.095 | 0.006 | 0.084...0.107 | 0.000 | * |
| Glaucoma (H40) | 0.349 | 0.071 | 0.210...0.488 | 0.000 | * |
| Essential (primary) hypertension (I10) | 0.006 | 0.002 | 0.001...0.010 | 0.022 | |
| Hypertensive heart disease (I11) | 0.017 | 0.002 | 0.013...0.021 | 0.000 | * |
| Angina pectoris (I20) | 0.015 | 0.005 | 0.005...0.024 | 0.002 | |
| Chronic ischaemic heart disease (I25) | 0.035 | 0.007 | 0.022...0.049 | 0.000 | * |
| Atrial fibrillation and flutter (I48) | -0.011 | 0.005 | -0.02...-0.002 | 0.020 | |
| Other cardiac arrhythmias (I49) | -0.012 | 0.007 | -0.025...0.001 | 0.075 | |
| Heart failure (I50) | 0.014 | 0.004 | 0.006...0.023 | 0.001 | |
| Other cerebrovascular diseases (I67) | -0.049 | 0.016 | -0.081...-0.018 | 0.002 | |
| Atherosclerosis (I70) | 0.005 | 0.006 | -0.007...0.017 | 0.414 | |
| Other chronic obstructive pulmonary disease (J44) | 0.037 | 0.008 | 0.021...0.054 | 0.000 | * |
| Asthma (J45) | 0.061 | 0.011 | 0.039...0.082 | 0.000 | * |
| Gastritis and duodenitis (K29) | -0.008 | 0.007 | -0.022...0.006 | 0.270 | |
| Functional dyspepsia (K30) | -0.050 | 0.008 | -0.065...-0.036 | 0.000 | * |
| Ulcerative colitis (K51) | 0.050 | 0.018 | 0.014...0.086 | 0.006 | |
| Chronic hepatitis, not elsewhere classified (K73) | 0.059 | 0.023 | 0.014...0.104 | 0.011 | |

| | | | | | |
|---|---|---|---|---|---|
| Fibrosis and cirrhosis of liver (K74) | 0.113 | 0.019 | 0.075...0.151 | 0.000 | * |
| Cholelithiasis (K80) | 0.053 | 0.027 | 0.000...0.105 | 0.048 | |
| Other diseases of pancreas (K86) | -0.002 | 0.007 | -0.015...0.011 | 0.748 | |
| Seropositive rheumatoid arthritis (M05) | 0.025 | 0.007 | 0.011...0.039 | 0.000 | * |
| Other rheumatoid arthritis (M06) | 0.024 | 0.007 | 0.010...0.038 | 0.001 | * |
| Psoriatic and enteropathic arthropathies (M07) | 0.016 | 0.012 | -0.009...0.040 | 0.206 | |
| Gout (M10) | -0.056 | 0.018 | -0.091...-0.021 | 0.002 | |
| Other inflammatory spondylopathies (M46) | -0.033 | 0.016 | -0.066...-0.001 | 0.043 | |
| Osteoporosis with pathological fracture (M80) | 0.086 | 0.019 | 0.049...0.123 | 0.000 | * |
| Osteoporosis without pathological fracture (M81) | 0.060 | 0.019 | 0.023...0.097 | 0.001 | |
| Hyperplasia of prostate (N40) | 0.018 | 0.006 | 0.006...0.030 | 0.004 | |
| Inflammatory diseases of prostate (N41) | -0.027 | 0.013 | -0.052...-0.002 | 0.037 | |
| Failure and rejection of transplanted organs and tissues (T86) | 0.099 | 0.022 | 0.057...0.142 | 0.000 | * |
| Transplanted organ and tissue status (Z94) | 0.077 | 0.024 | 0.030...0.123 | 0.001 | |
| Presence of cardiac and vascular implants and grafts (Z95) | 0.061 | 0.006 | 0.050...0.072 | 0.000 | * |
| Ingredient x Disease | | | | | |
| Amlodipine:Hypertensive heart disease (I11) | -0.006 | 0.004 | -0.015...0.003 | 0.186 | |
| Atenolol:Hypertensive heart disease (I11) | 0.026 | 0.018 | -0.010...0.062 | 0.153 | |
| Bisoprolol:Hypertensive heart disease (I11) | -0.020 | 0.008 | -0.035...-0.005 | 0.009 | |
| Candesartan:Hypertensive heart disease (I11) | 0.013 | 0.012 | -0.010...0.035 | 0.274 | |
| Carvedilol:Hypertensive heart disease (I11) | -0.042 | 0.014 | -0.069...-0.014 | 0.003 | |
| Doxazosin:Hypertensive heart disease (I11) | -0.026 | 0.012 | -0.050...-0.003 | 0.026 | |
| Enalapril:Hypertensive heart disease (I11) | 0.002 | 0.007 | -0.012...0.015 | 0.786 | |
| Felodipine:Hypertensive heart disease (I11) | 0.022 | 0.020 | -0.016...0.061 | 0.260 | |
| Fosinopril:Hypertensive heart disease (I11) | 0.016 | 0.009 | -0.002...0.034 | 0.073 | |
| Hydrochlorothiazide:Hypertensive heart disease (I11) | 0.020 | 0.005 | 0.011...0.030 | 0.000 | * |
| Indapamide:Hypertensive heart disease (I11) | -0.001 | 0.006 | -0.013...0.010 | 0.794 | |
| Lacidipine:Hypertensive heart disease (I11) | -0.017 | 0.017 | -0.050...0.016 | 0.314 | |
| Lercanidipine:Hypertensive heart disease (I11) | -0.010 | 0.012 | -0.032...0.013 | 0.396 | |
| Lisinopril:Hypertensive heart disease (I11) | -0.002 | 0.024 | -0.050...0.045 | 0.923 | |
| Losartan:Hypertensive heart disease (I11) | -0.013 | 0.012 | -0.037...0.012 | 0.311 | |
| Moxonidine:Hypertensive heart disease (I11) | -0.039 | 0.011 | -0.060...-0.018 | 0.000 | * |
| Nebivolol:Hypertensive heart disease (I11) | -0.003 | 0.006 | -0.015...0.008 | 0.597 | |
| Nifedipine:Hypertensive heart disease (I11) | -0.030 | 0.017 | -0.064...0.004 | 0.083 | |
| Nitrendipine:Hypertensive heart disease (I11) | 0.007 | 0.011 | -0.014...0.028 | 0.541 | |
| Olmesartan:Hypertensive heart disease (I11) | -0.006 | 0.009 | -0.025...0.012 | 0.517 | |
| Perindopril:Hypertensive heart disease (I11) | -0.002 | 0.006 | -0.013...0.009 | 0.765 | |
| Propranolol:Hypertensive heart disease (I11) | 0.003 | 0.015 | -0.027...0.033 | 0.856 | |
| Ramipril:Hypertensive heart disease (I11) | -0.003 | 0.004 | -0.011...0.006 | 0.513 | |
| Telmisartan:Hypertensive heart disease (I11) | -0.010 | 0.005 | -0.021...0.000 | 0.057 | |
| Trandolapril:Hypertensive heart disease (I11) | -0.014 | 0.025 | -0.062...0.035 | 0.584 | |
| Trimetazidine dihydrochloride:Hypertensive heart disease (I11) | -0.028 | 0.006 | -0.040...-0.017 | 0.000 | * |
| Valsartan:Hypertensive heart disease (I11) | -0.012 | 0.013 | -0.037...0.012 | 0.324 | |
| Verapamil:Hypertensive heart disease (I11) | -0.006 | 0.009 | -0.024...0.011 | 0.475 | |
| Amlodipine:Essential (primary) hypertension (I10) | 0.008 | 0.004 | -0.001...0.017 | 0.073 | |
| Atenolol:Essential (primary) hypertension (I10) | 0.049 | 0.019 | 0.013...0.086 | 0.008 | |
| Candesartan:Essential (primary) hypertension (I10) | 0.017 | 0.012 | -0.006...0.040 | 0.147 | |
| Enalapril:Essential (primary) hypertension (I10) | 0.006 | 0.007 | -0.008...0.020 | 0.380 | |
| Felodipine:Essential (primary) hypertension (I10) | 0.028 | 0.020 | -0.010...0.067 | 0.152 | |
| Fosinopril:Essential (primary) hypertension (I10) | 0.013 | 0.009 | -0.005...0.031 | 0.171 | |

| | | | | | |
|---|---|---|---|---|---|
| Hydrochlorothiazide:Essential (primary) hypertension (I10) | 0.025 | 0.005 | 0.015...0.035 | 0.000 | * |
| Indapamide:Essential (primary) hypertension (I10) | 0.005 | 0.006 | -0.007...0.016 | 0.435 | |
| Lacidipine:Essential (primary) hypertension (I10) | -0.001 | 0.017 | -0.034...0.032 | 0.949 | |
| Lercanidipine:Essential (primary) hypertension (I10) | -0.002 | 0.012 | -0.025...0.021 | 0.843 | |
| Lisinopril:Essential (primary) hypertension (I10) | 0.021 | 0.024 | -0.026...0.068 | 0.381 | |
| Losartan:Essential (primary) hypertension (I10) | -0.009 | 0.012 | -0.033...0.015 | 0.462 | |
| Nebivolol:Essential (primary) hypertension (I10) | -0.001 | 0.006 | -0.013...0.011 | 0.882 | |
| Nifedipine:Essential (primary) hypertension (I10) | -0.042 | 0.018 | -0.076...-0.007 | 0.017 | |
| Nitrendipine:Essential (primary) hypertension (I10) | 0.007 | 0.011 | -0.014...0.029 | 0.501 | |
| Olmesartan:Essential (primary) hypertension (I10) | -0.005 | 0.009 | -0.023...0.014 | 0.618 | |
| Perindopril:Essential (primary) hypertension (I10) | 0.004 | 0.006 | -0.007...0.015 | 0.458 | |
| Propranolol:Essential (primary) hypertension (I10) | -0.020 | 0.015 | -0.049...0.008 | 0.162 | |
| Ramipril:Essential (primary) hypertension (I10) | 0.001 | 0.005 | -0.007...0.010 | 0.743 | |
| Telmisartan:Essential (primary) hypertension (I10) | -0.001 | 0.005 | -0.011...0.010 | 0.926 | |
| Trandolapril:Essential (primary) hypertension (I10) | -0.003 | 0.025 | -0.051...0.046 | 0.918 | |
| Valsartan:Essential (primary) hypertension (I10) | -0.014 | 0.013 | -0.039...0.011 | 0.267 | |
| Verapamil:Essential (primary) hypertension (I10) | 0.007 | 0.010 | -0.013...0.027 | 0.475 | |
| Empagliflozin:Type 2 diabetes mellitus (E11) | -0.007 | 0.126 | -0.254...0.241 | 0.957 | |
| Gliclazide:Type 2 diabetes mellitus (E11) | -0.193 | 0.104 | -0.396...0.011 | 0.063 | |
| Glimepiride:Type 2 diabetes mellitus (E11) | -0.190 | 0.108 | -0.401...0.021 | 0.078 | |
| Insulin aspart:Type 2 diabetes mellitus (E11) | -0.230 | 0.102 | -0.429...-0.031 | 0.023 | |
| Insulin detemir:Type 2 diabetes mellitus (E11) | -0.204 | 0.102 | -0.403...-0.005 | 0.045 | |
| Insulin glargine:Type 2 diabetes mellitus (E11) | -0.185 | 0.102 | -0.384...0.015 | 0.069 | |
| Insulin glulisine, human:Type 2 diabetes mellitus (E11) | -0.251 | 0.103 | -0.452...-0.050 | 0.014 | |
| Insulin lispro:Type 2 diabetes mellitus (E11) | -0.155 | 0.102 | -0.355...0.045 | 0.130 | |
| Linagliptin:Type 2 diabetes mellitus (E11) | -0.081 | 0.119 | -0.314...0.152 | 0.497 | |
| Liraglutide:Type 2 diabetes mellitus (E11) | -0.341 | 0.117 | -0.570...-0.111 | 0.004 | |
| Metformin:Type 2 diabetes mellitus (E11) | -0.168 | 0.101 | -0.367...0.030 | 0.097 | |
| Saxagliptin:Type 2 diabetes mellitus (E11) | -0.077 | 0.125 | -0.322...0.167 | 0.535 | |
| Sitagliptin:Type 2 diabetes mellitus (E11) | -0.099 | 0.110 | -0.314...0.116 | 0.367 | |
| Betaxolol:Glaucoma (H40) | -0.171 | 0.102 | -0.371...0.029 | 0.093 | |
| Bimatoprost:Glaucoma (H40) | -0.421 | 0.229 | -0.869...0.028 | 0.066 | |
| Brinzolamide:Glaucoma (H40) | 0.012 | 0.113 | -0.210...0.234 | 0.915 | |
| Dorzolamide:Glaucoma (H40) | -0.272 | 0.080 | -0.429...-0.115 | 0.001 | * |
| Latanoprost:Glaucoma (H40) | -0.319 | 0.088 | -0.492...-0.146 | 0.000 | * |
| Timolol:Glaucoma (H40) | -0.219 | 0.077 | -0.370...-0.068 | 0.005 | |
| Atorvastatin:Disorders of lipoprotein metabolism and other lipidaemias (E78) | -0.011 | 0.005 | -0.021...-0.001 | 0.029 | |
| Fluvastatin:Disorders of lipoprotein metabolism and other lipidaemias (E78) | 0.034 | 0.017 | 0.002...0.067 | 0.038 | |
| Pravastatin:Disorders of lipoprotein metabolism and other lipidaemias (E78) | 0.017 | 0.020 | -0.021...0.056 | 0.377 | |
| Rosuvastatin:Disorders of lipoprotein metabolism and other lipidaemias (E78) | -0.012 | 0.005 | -0.022...-0.002 | 0.019 | |
| Carvedilol:Heart failure (I50) | 0.012 | 0.013 | -0.015...0.038 | 0.384 | |
| Digoxin:Heart failure (I50) | -0.009 | 0.011 | -0.030...0.013 | 0.432 | |
| Furosemide:Heart failure (I50) | -0.018 | 0.009 | -0.036...0.001 | 0.058 | |
| Spironolactone:Heart failure (I50) | -0.002 | 0.007 | -0.016...0.012 | 0.769 | |
| Amiodarone:Atrial fibrillation and flutter (I48) | 0.025 | 0.010 | 0.005...0.044 | 0.015 | |
| Apixaban:Atrial fibrillation and flutter (I48) | 0.017 | 0.013 | -0.007...0.042 | 0.167 | |
| Dabigatran:Atrial fibrillation and flutter (I48) | 0.085 | 0.012 | 0.061...0.109 | 0.000 | * |
| Propafenone:Atrial fibrillation and flutter (I48) | 0.047 | 0.009 | 0.029...0.065 | 0.000 | * |

| | | | | | |
|---|---|---|---|---|---|
| Rivaroxaban:Atrial fibrillation and flutter (I48) | 0.037 | 0.008 | 0.022...0.052 | 0.000 | * |
| Sotalol:Atrial fibrillation and flutter (I48) | 0.026 | 0.011 | 0.003...0.048 | 0.024 | |
| Verapamil:Atrial fibrillation and flutter (I48) | 0.029 | 0.012 | 0.005...0.053 | 0.016 | |
| Albuterol:Asthma (J45) | 0.045 | 0.012 | 0.021...0.069 | 0.000 | * |
| Formoterol:Asthma (J45) | -0.079 | 0.013 | -0.105...-0.053 | 0.000 | * |
| Ipratropium:Asthma (J45) | -0.017 | 0.014 | -0.045...0.010 | 0.212 | |
| Montelukast:Asthma (J45) | 0.027 | 0.014 | -0.001...0.056 | 0.057 | |
| Alfuzosin:Hyperplasia of prostate (N40) | -0.004 | 0.013 | -0.030...0.023 | 0.786 | |
| Doxazosin:Hyperplasia of prostate (N40) | -0.007 | 0.012 | -0.032...0.017 | 0.571 | |
| Dutasteride:Hyperplasia of prostate (N40) | -0.009 | 0.015 | -0.040...0.021 | 0.542 | |
| Agomelatine:F32 | -0.017 | 0.023 | -0.062...0.028 | 0.457 | |
| Bupropion:Depressive episode (F32) | 0.001 | 0.023 | -0.044...0.046 | 0.980 | |
| Citalopram:Depressive episode (F32) | 0.017 | 0.022 | -0.026...0.060 | 0.434 | |
| Duloxetine:Depressive episode (F32) | 0.018 | 0.022 | -0.024...0.061 | 0.400 | |
| Escitalopram:Depressive episode (F32) | -0.004 | 0.020 | -0.044...0.035 | 0.833 | |
| Fluoxetine:Depressive episode (F32) | 0.022 | 0.021 | -0.019...0.063 | 0.297 | |
| Flupenthixol:Depressive episode (F32) | 0.002 | 0.029 | -0.054...0.058 | 0.951 | |
| Mirtazapine:Depressive episode (F32) | 0.024 | 0.021 | -0.017...0.065 | 0.254 | |
| Nortriptyline:Depressive episode (F32) | -0.001 | 0.027 | -0.055...0.053 | 0.965 | |
| Paroxetine:Depressive episode (F32) | 0.008 | 0.022 | -0.034...0.051 | 0.709 | |
| Sertraline:Depressive episode (F32) | 0.014 | 0.021 | -0.027...0.055 | 0.497 | |
| Tianeptine:Depressive episode (F32) | -0.010 | 0.021 | -0.051...0.032 | 0.653 | |
| Venlafaxine:Depressive episode (F32) | 0.003 | 0.021 | -0.039...0.045 | 0.889 | |
| Allopurinol:Gout (M10) | 0.052 | 0.019 | 0.014...0.089 | 0.007 | |

[1]Statistically significant after Bonferroni correction

**Supplementary Table 5. The effect of person-specific medication adherence on cause-specific hospitalisation in Cox proportional hazards model.**

| Disease (ICD-10 code) | N of events | Risk population | Hazard ratio | 95% CI | p-value | Sig[1] |
|---|---|---|---|---|---|---|
| Myocardial infraction (I21-I23) | 508 | 47 064 | 0.43 | 0.19…0.99 | 0.046 | |
| Cardiac arrhythmias (I46-I49) | 661 | 46 671 | 0.76 | 0.37…1.59 | 0.470 | |
| Heart failure (I50) | 320 | 47 596 | 0.63 | 0.22…1.83 | 0.398 | |
| Cerebral ischemia/stroke (I60-I64, I69, G45) | 882 | 46 536 | 0.44 | 0.23…0.82 | 0.009 | |
| Injuries (S00-S99) | 1122 | 46 163 | 0.59 | 0.34…1.03 | 0.061 | |
| Hip fracture (S72) | 367 | 47 535 | 1.03 | 0.37…2.81 | 0.961 | |
| Pneumonia (J12-J18) | 576 | 41 106 | 1.54 | 0.69…3.46 | 0.295 | |
| Urinary tract infection (N10, N12, N15, N30) | 114 | 47 759 | 0.40 | 0.07…2.24 | 0.294 | |
| Gastrointestinal bleeding (K25-K28) | 166 | 47 676 | 1.65 | 0.36…7.45 | 0.517 | |

[1] Bonferroni adjusted p-value: 0.006

**Supplementary Table 6. The effect of person-specific medication adherence on cause-specific incidence in Cox proportional hazards model.**

| Cause | Cases | Risk population | Hazard ratio | 95%CI | p-valueSig[1] |
|---|---|---|---|---|---|
| Disorders of thyroid gland (E00-E07) | 1 442 | 39 772 | 0.59 | 0.36…0.95 | 0.031 |
| Diabetes mellitus (E10-E14) | 1 191 | 39 781 | 1.44 | 0.83…2.51 | 0.193 |
| Disorders of lipoprotein metabolism and other lipidaemias (E78) | 2 929 | 31 728 | 0.48 | 0.34…0.68 | < 0.001* |
| Depression (F32, F33) | 1 543 | 38 595 | 0.76 | 0.47…1.22 | 0.255 |
| Neurotic disorders (F40-F48) | 2 245 | 36 492 | 0.71 | 0.48…1.06 | 0.093 |
| Anemia (D50-D64) | 2 728 | 42 485 | 0.96 | 0.67…1.38 | 0.821 |
| Ischemic heart disease (I20-I25) | 2 252 | 37 801 | 0.40 | 0.27…0.59 | < 0.001* |
| Cardiac valve disorders (I34-I37) | 543 | 46 561 | 0.82 | 0.36…1.85 | 0.630 |
| Cardic arrhythmias (I46-I49) | 3 115 | 37 304 | 0.66 | 0.47…0.92 | 0.014 |
| Heart failure (I50) | 2 603 | 38 783 | 0.85 | 0.59…1.23 | 0.395 |
| Cerebral ischemia/stroke (I60-I64, I69, G45) | 1 607 | 43 828 | 0.45 | 0.29…0.72 | < 0.001* |
| Atherosclerosis/peripheral arterial occlusive disease (I65-I67, I70, I73) | 1 898 | 41 970 | 0.27 | 0.18….0.41 | < 0.001* |
| Asthma and chronic obstructive bronchitis (J43-J47) | 1 677 | 40 123 | 0.32 | 0.20…0.50 | < 0.001* |
| Chronic cholecystitis/ gallstones (K80-K81) | 1 593 | 44 003 | 0.42 | 0.26…0.66 | < 0.001* |
| Osteoporosis (M80-M82) | 985 | 45 580 | 0.30 | 0.17…0.54 | < 0.001* |
| Renal failure (N17-N19) | 1 671 | 46 059 | 0.82 | 0.51…1.30 | 0.394 |
| Gout (M10) | 1 608 | 44 137 | 1.18 | 0.74…1.90 | 0.488 |
| Diseases of liver (K70-K77) | 1 094 | 45 678 | 0.27 | 0.15…0.47 | < 0.001* |
| Parkinson's disease (G20-G22) | 251 | 47 271 | 2.05 | 0.59…7.08 | 0.257 |
| Diseases of stomach (K20-K31) | 4 214 | 28 702 | 0.58 | 0.44…0.78 | < 0.001* |
| Insomnia (G47, F51) | 3 087 | 35 667 | 0.45 | 0.32…0.63 | < 0.001* |
| Dementia (F00-F03, G30, G31) | 988 | 46 314 | 0.46 | 0.25…0.84 | 0.011 |
| Varicose veins of lower extremities (I83, I87) | 1 496 | 43 891 | 0.30 | 0.19…0.49 | < 0.001* |
| Migraine/chronic headache (G43-G44) | 1 137 | 44 205 | 0.36 | 0.21…0.64 | < 0.001* |
| Dizziness (H81-H82, R42) | 2 432 | 41 620 | 0.37 | 0.25…0.54 | < 0.001* |
| Severe hearing loss (H90-H91) | 2 202 | 43 073 | 0.46 | 0.31…0.68 | < 0.001* |
| Aneurysm, thrombosis, embolism (I71, I72, I74, I80, I81, I82, I26) | 1 520 | 44 885 | 0.32 | 0.20…0.51 | < 0.001* |
| Injuries (S00-S99) | 5 634 | 30 497 | 0.49 | 0.38…0.62 | < 0.001* |
| Hip fracture (S72) | 439 | 47 434 | 0.78 | 0.31…1.93 | 0.587 |
| Pneumonia (J12-J18) | 2 875 | 43 342 | 0.77 | 0.54…1.09 | 0.134 |
| Urinary tract infection (N10, N12, N15, N30) | 2 649 | 39 121 | 0.67 | 0.47…0.97 | 0.034 |
| Gastrointestinal bleeding (K25-K28) | 1 060 | 44 147 | 0.34 | 0.19…0.60 | < 0.001* |

[1] Bonferroni adjusted p-value: 0.002

**Supplementary Table 7.** The number of cases included into the Cox proportional hazards model to estimate incidence and hospitalisation for selected diseases.

| Cause | ICD-10 codes | Incidence | | Hospitalisation | |
|---|---|---|---|---|---|
| | | Cases | Risk population | Cases | Risk population |
| Disorders of thyroid gland | E00-E07 | 1442 | 39772 | | |
| Diabetes mellitus | E10-E14 | 1191 | 39781 | | |
| Disorders of lipoprotein metabolism and other lipidaemias | E78 | 2929 | 31728 | | |
| Depression | F32, F33 | 1543 | 38595 | | |
| Neurotic disorders | F40-F48 | 2245 | 36492 | | |
| Anemia | D50-D64 | 2728 | 42485 | | |
| Ischemic heart disease | I20-I25 | 2252 | 37801 | | |
| Myocardial infraction | I21-I23 | | | 508 | 47064 |
| Cardiac valve disorders | I34-I37 | 543 | 46561 | | |
| Cardic arrhythmias | I46-I49 | 3115 | 37304 | 661 | 46671 |
| Heart failure | I50 | 2603 | 38783 | 320 | 47596 |
| Cerebral ischemia/stroke | I60-I64, I69, G45 | 1607 | 43828 | 882 | 46536 |
| Atherosclerosis/peripheral arterial occlusive disease | I65-I67, I70, I73 | 1898 | 41970 | | |
| Asthma and chronic obstructive bronchitis | J43-J47 | 1677 | 40123 | | |
| Chronic cholecystitis/ gallstones | K80-K81 | 1593 | 44003 | | |
| Osteoporosis | M80-M82 | 985 | 45580 | | |
| Renal failure | N17-N19 | 1671 | 46059 | | |
| Gout | M10 | 1608 | 44137 | | |
| Diseases of liver | K70-K77 | 1094 | 45678 | | |
| Parkinson's disease | G20-G22 | 251 | 47271 | | |
| Diseases of stomach | K20-K31 | 4214 | 28702 | | |
| Insomnia | G47, F51 | 3087 | 35667 | | |
| Dementia | F00-F03, G30, G31 | 988 | 46314 | | |
| Varicose veins of lower extremities | I83, I87 | 1496 | 43891 | | |
| Migrane/chornic headache | G43-G44 | 1137 | 44205 | | |
| Dizziness | H81-H82, R42 | 2432 | 41620 | | |
| Severe hearing loss | H90-H91 | 2202 | 43073 | | |
| Aneurysm, thrombosis, embolism | I71, I72, I74, I80, I81, I82, I26 | 1520 | 44885 | | |
| Injuries | S00-S99 | 5634 | 30497 | 1122 | 46163 |
| Hip fracture | S72 | 439 | 47434 | 367 | 47535 |
| Pneumonia | J12-J18 | 2875 | 43342 | 576 | 47106 |
| Urinary tract infection | N10, N12, N15, N30 | 2649 | 39121 | 114 | 47759 |
| Gastrointestinal bleeding | K25-K28 | 1060 | 44147 | 166 | 47676 |
